# Use of Federated Learning for validating and updating privacy-preserving decentralized multi-study prognostic models in Traumatic Brain Injury

**DOI:** 10.64898/2026.07.30.26359337

**Authors:** Abel Torres-Espin, Justin C. Wong, H. E. Hinson, Thomas B. Kuipers, Bart P.T. Hoekstra, Sonia Jain, Xiaoying Sun, John K. Yue, Dana Pisică, Ana Mikolic, Hester F. Lingsma, Amy J. Markowitz, Adam R. Ferguson, David K. Menon, Andrew I. R. Maas, Ewout W. Steyerberg, Geoffrey T. Manley, Patrick J. Belton

## Abstract

Developing modern clinical prediction models (CPMs) and advanced analytics requires large datasets, often necessitating data from different studies. Privacy regulations may hinder data sharing, especially across countries. Decentralized federated data infrastructures, where data remain in their original location and analyses are run only in a shared, secure environment, may address these challenges. We implemented a privacy-preserving federated learning (FL) infrastructure and evaluated and updated the IMPACT prognostic models for traumatic brain injury (TBI) using 2 studies. A multi-continental federated infrastructure was established between 2 large-scale studies (TRACK-TBI from the United States and CENTER-TBI from Europe and Israel). Three IMPACT prognostic models for post-TBI 6-month mortality and unfavorable outcomes were evaluated, followed by model updates through 2 FL approaches trained across the TRACK-TBI and CENTER-TBI studies. Internal validation, external cross-validation, and sub-study validations were performed. CPMs were evaluated for discrimination and calibration. The federated cohort included 1616 participants (TRACK-TBI: n=441, CENTER-TBI: n=1175). Both FL performed well, with comparable coefficient estimates, AUCs (area under the receiver operating characteristics curve) between 0.77-0.88, and calibrated probabilities. Compared to the original IMPACT and single-study models, both federated models presented similar discrimination (AUC), were well-calibrated, were more efficient (higher precision), and reduced the impact of missing data in model estimation. FL is feasible for privacy-preserving development and evaluation of CPMs, and can enable validation and updating across large, virtually analyzed datasets while overcoming regulatory constraints on data combination. Federated infrastructures can facilitate global collaboration to advance data-hungry analytical methods, such as artificial intelligence.

## INTRODUCTION

Traumatic brain injury (TBI) is a leading cause of death and long-term disability worldwide, posing a considerable public health problem.^1^ Early, accurate prognostication in TBI is essential to support clinical decision-making, inform patients and relatives, and advance research.^1^ The IMPACT (International Mission for Prognosis and Analysis of Clinical Trials) prognostic models comprise a set of 3 models with an increasing number of predictors **(Table 1)**, designed to predict 2 different 6-month outcomes: mortality, and unfavorable outcome measured by Glasgow Outcome Scale (GOS)^2^ score 1-3 (death to severe disability). The models were originally developed from international datasets of moderate-severe TBI patients (Glasgow Coma Scale [GCS] ≤12), from 8 randomized controlled trials and 3 observational studies conducted between 1984 and 1997.^3^ Since then, the models have been extensively used and externally validated.^4–8^ Recent validation studies suggest the need to re-evaluate and update the models to incorporate contemporary changes in TBI management and maintain currentness and generalizability.^6,8^ Modern efforts to develop, evaluate, and update clinical prediction models across countries are challenged by global data privacy regulations for international sharing of human subject data.^9,10^

**Table 1.** Set of predictors for each tier of the original IMPACT TBI prognostic model.

| *Model* | *Predictor* | *Description* | *Type* |
| --- | --- | --- | --- |
| *Base* | Age | Age at injury in years | Continuous |
|  | GCS motor score | GCS motor score (None (1), Extension (2), Abnormal flexion (3), Normal flexion (4), and Localizes/obeys (5/6)) | Categorical, Reference '5' |
|  | Pupils | Pupillary reactivity (both reactive, one reactive, none reactive) | Categorical, reference 'both' |
| *Extended (Base +)* | Marshall | Marshall CT score (I, II, III/IV, V/VI) | Categorical, reference 'II' |
|  | tSAH | Traumatic subarachnoid hemorrhage on initial head CT (Yes, No) | Binary, reference 'No' |
|  | EDH | Epidural hematoma on initial head CT (Yes, No) | Binary, reference 'No' |
|  | Hypoxia | Presentation of hypoxia (Yes, No) | Binary, reference 'No' |
|  | Hypotension | Presentation of hypotension (Yes, No) | Binary, reference 'No' |
| *Lab (Extended +)* | Glucose | Glucose levels (mmol/l) | Continuous |
|  | Hemoglobin | Hemoglobin levels (g/dl) | Continuous |

Federated learning (FL), an approach that allows training statistical and machine learning models across multiple decentralized computers, is an attractive alternative, as it brings the analysis to the data, keeping the data local rather than sharing or combining data in a centralized analysis environment. FL overcomes some of the challenges of multi-continental data sharing, while protecting patient privacy.^11,12^ Previous research has demonstrated the application of FL in different medical contexts.^12–15^ Our first objective was to assess the feasibility of a federated infrastructure with a decentralized analytical workflow for prediction model validation and updating in TBI across continents. The second objective was to assess whether a global IMPACT model, that is, a model applicable to any given study with similar cohort characteristics, remains relevant and whether it can be updated using modern datasets through FL. We implemented a distributed computer network that enables privacy-preserving decentralized analysis and modeling across 2 recently completed, multi-institutional, prospectively enrolling observational studies: the Transforming Research and Clinical Knowledge in Traumatic Brain Injury Study (TRACK-TBI)^16^ from 18 level 1 trauma centers in the United States (US) and the Collaborative European NeuroTrauma Effectiveness Research in Traumatic Brain Injury Study (CENTER-TBI)^17^ from 68 centers in Europe and Israel **(Figure 1)**.

**Figure 1.**
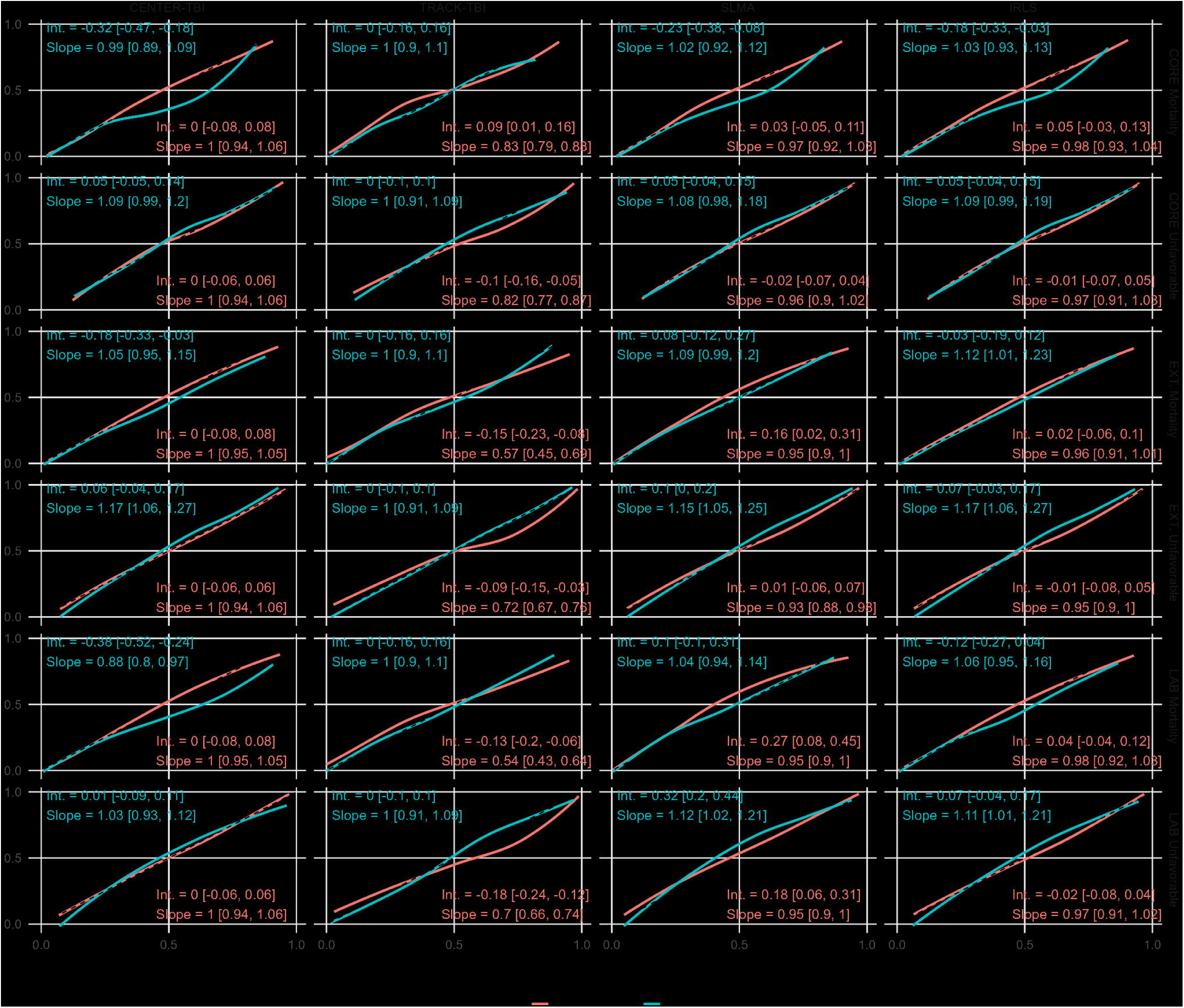
Privacy-preserving federated system between CENTER-TBI and TRACK-TBI. (a) The distributed federated system has 2 nodes, one in Leiden, Netherlands (CENTER-TBI), and 1 in San Francisco, United States (TRACK-TBI). Each node has a private local server with study data and computing capabilities protected by a firewall. The analysis and modeling code is sent over the internet through a federated client (DataSHIELD) simultaneously to each node. The code request is evaluated locally (eg, harmonization transformation, summary statistics, modeling functions) and non-disclosive results are returned to the user managed by the federated client. We used 2 different federated methods to fit prediction models after harmonization and data preparation: (b) a 2-step study level meta-analysis where an intermediate model is first fitted independently for each study, followed by pooling the estimates; and (c) where a global model is iteratively fitted by sharing intermediate estimates with the nodes at each iteration and updating them. This is conducted until estimates do not change, at which point the model has converged. After models are fitted, we performed subset validation by evaluating prediction performance in each study independently.

## MATERIALS AND METHODS

### Study Design, Setting and Data

Our federated data infrastructure, a distributed computing network,^11,12^ enables privacy-preserving decentralized analysis and model training across the 2 databases. We employed FL to validate and update prediction models in decentralized networks. From each study, a subset of individuals meeting inclusion criteria of the original cohorts for each model was selected as previously used for single-database validation studies.^5,6^ In brief, TBI patients with a presenting GCS ≤12, age ≥14 years at time of injury, and available GOS Extended (GOSE)^18^ score at 6-months post-injury were included in the analyses. The Transparent Reporting of a multivariable Prediction model for individual Prognosis Or Diagnosis (TRIPOD) reporting guidelines were followed.^19^ TRACK-TBI and CENTER-TBI data were collected under ethical approval (see below).

The TRACK-TBI Study was approved by institutional review boards (IRBs) at each study site (Baylor College of Medicine, Massachusetts General Hospital/Spaulding Rehabilitation Hospital, University of California, San Francisco, University of Cincinnati, University of Maryland, University of Miami, University of Pittsburgh, University of Texas at Austin, University of Texas Southwestern, University of Washington, Virginia Commonwealth University, University of Pennsylvania, Emory University, Medical College of Wisconsin, University of Utah, Indiana University, Hennepin Healthcare, University of Colorado). The overall study received approval from the IRB of record (i.e., Ethics Reviewer) at the University of California, San Francisco (Protocol Number: 10-00111). Each participant (or their legally authorized representative) provided written informed consent.

The CENTER-TBI study (EC grant 602150) has been conducted in accordance with all relevant laws of the EU if directly applicable or of direct effect and all relevant laws of the country where the Recruiting sites were located, including but not limited to, the relevant privacy and data protection laws and regulations (the “Privacy Law”), the relevant laws and regulations on the use of human materials, and all relevant guidance relating to clinical studies from time to time in force including, but not limited to, the ICH Harmonised Tripartite Guideline for Good Clinical Practice (CPMP/ICH/135/95) (“ICH GCP”) and the World Medical Association Declaration of Helsinki entitled “Ethical Principles for Medical Research Involving Human Subjects”. Informed Consent by the patients and/or the legal representative/next of kin was obtained, accordingly to the local legislations, for all patients recruited in the Core Dataset of CENTER-TBI and documented in the e-CRF. Ethical approval was obtained for each recruiting sites. The list of sites, Ethical Committees, approval numbers and approval dates can be found here: https://www.center-tbi.eu/project/ethical-approval

### Federated Infrastructure

The distributed network comprises 2 server nodes, one at Leiden, the Netherlands (CENTER-TBI database) and 1 at the University of California, San Francisco, California, US (TRACK-TBI database), under respective firewalls and network security **(Figure 1a)**. Each node uses Opal^20^ as the main data warehouse and an R server, allowing for local computing. We used the DataSHIELD tools^21,22^ and framework for federated computing, including data preparation, harmonization, and modeling. All coding for analysis and modeling was prepared in the local client computer of an analyst in R and DataSHIELD, which sends code simultaneously to each node through application programming interface (API) calls, triggering the computing in each node’s R server and returning aggregated results if compliant with privacy-preserving policies.

### Data Preparation

Cohort selection and data harmonization are performed through the federated infrastructure; analysts have no access to either dataset on their local machines. Five multiple imputations were conducted to independently impute missing data on the predictors through multivariate imputation by chained equation (mice)^23^ on each server side. No outcomes were imputed, and individuals without outcomes were excluded from analysis. Each complete imputed dataset was used for modeling and evaluation. Parameter estimates and performance metrics were pooled as specified below. Procedures were identical on both servers.

### Prediction Models and Federated Learning

Validation of the original IMPACT prediction models^3^ was previously reported for both TRACK-TBI and CENTER-TBI.^5,6^ As in those reports, we defined unfavorable outcome as GOSE = 1-4 and mortality as GOSE = 1. There are 3 different versions of the IMPACT model, with increasing numbers of predictors (core, extended, and laboratory, **Table 1**). The IMPACT models are specified as logistic regressions with a logit link function. In the context of FL, logistic regression parameters were estimated using 2 different approaches **(Figure 1)**: a 2-stage meta-analysis approach and a co-learning approach^24–26^. The 2-stage FL meta-analysis **(Figure 1b)** consists of first fitting the logistic model in each node to convergence and pooling the model estimates, and in the second step using a random-effects meta-analysis model estimated through maximum likelihood. Referred to as study-level meta-analysis (SLMA), it is achieved using the *ds.glmSLMA* R function from the dsBaseClient R package. The co-learning FL estimates **(Figure 1c)** are obtained using iterative reweighting least squares (IRLS), as implemented in the *ds.glm* R function from the dsBaseClient R package. The IRLS algorithm iteratively fits the model in the local node of the federated network and shares information about the model estimates in each iteration with the other nodes; thus, raw data are never shared. The algorithm continues until convergence. Further, the coefficient estimates for each independent imputed dataset were pooled using Rubin’s rules^27^ with a custom adaptation of the implementation in the mice R package^23^ to the federated data privacy-preserving scenario. The relative efficiency due to missingness (RE) and the fraction of missing information (FMI) were calculated to measure the impact of missing imputation in the estimation.^28^

### Model Evaluation

We first validated the original IMPACT models’ coefficients in the CENTER-TBI and TRACK-TBI studies. Then, models were updated by independently re-estimating coefficients on the CENTER-TBI and TRACK-TBI studies and by pooling cohorts through SLMA and IRLS federated methods. The single-study models (i.e., CENTER-TBI and TRACK-TBI) were validated in their training data and by cross-evaluation (i.e., CENTER-TBI model evaluated in TRACK-TBI dataset, and *vice versa*). The latter effectively constitutes an external model validation over the federated network. The federated models based on the FL-pooled CENTER-TBI and TRACK-TBI data (SLMA and IRLS) were assessed by sub-evaluation (i.e., the model performance in each individual CENTER-TBI and TRACK-TBI datasets). Five different evaluations were conducted:

1. Model goodness-of-fit by the Nagelkerke pseudo-R^2^
2. Difference in coefficients, reflecting variations in model estimates.
3. Area under the operating receiving curve (AUC) as a metric of discrimination.
4. The Brier score (BS) is calculated 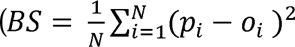, where *p_i_* and *O_i_*; are the predicted probability and observed outcome) to measure the average accuracy of probabilistic predictions as a metric of model calibration combined with discrimination. We report the Brier score scaled (BSs) 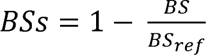, where *BS_ref_* is the BS for non-informative model prediction based on the prevalence), representing the percentage improvement in the BS compared with the reference model.
5. Calibration curves were constructed over the 0, 5, 10, 25, 50, 75, 90, 95, and 100 quantiles of the predicted probabilities. Calibration intercept and slope were calculated by fitting a logistic linear regression on the server side with the binary outcome (mortality or unfavorable outcome) as response and the logit of the predicted probabilities as predictor.

The AUCs, BS, and BSs over multiple imputations were pooled using meta-analysis through maximum likelihood estimation of heterogeneity implemented in the *rma* R function from the R metafor package.^29^

### Statistical Analysis

Differences between CENTER-TBI and TRACK-TBI cohorts **(eTable 1)** on predictors and outcomes were tested using a Fisher’s independence test for categorical and binary variables and a t-test for continuous variables. The pooled coefficient estimates between models were statistically compared by calculating the 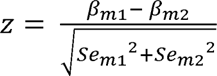 where *β* and *Se* are the coefficients and their standard errors to be compared, respectively. A 2-tailed p-value for the coefficient comparison was calculated using the standard normal probability density function, with a p-value < 0.05 considered statistically significant. Comparison of coefficients between models are shown in **eFigure 2**.

**Figure 2.**
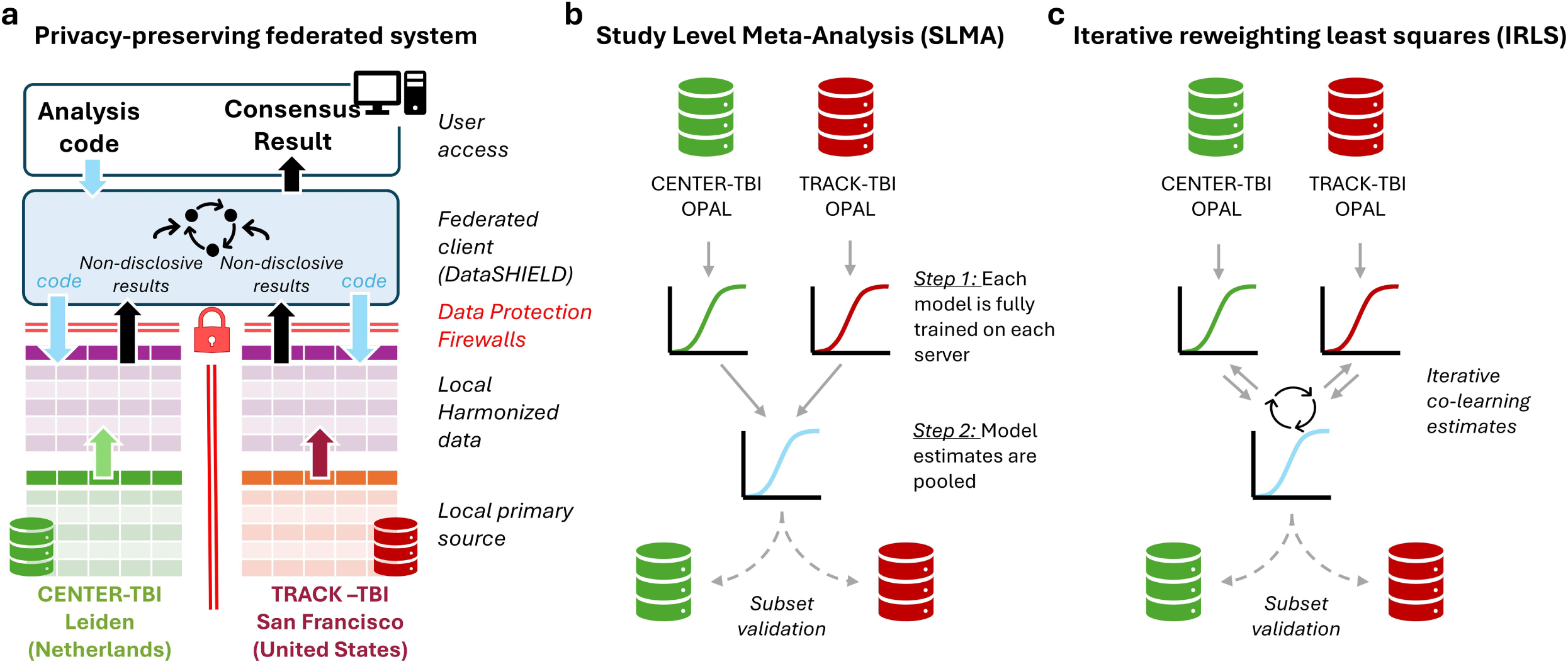
A calibration curve is shown for the average calibration curve across the 5 imputations. The ideal calibration is shown with a dotted black line. The estimated calibration intercept (Int.) and Slope together with 95% CI are shown for each validation dataset.

### Feasibility of FL

We considered the feasibility of performing FL through two outcomes. First, the capacity to implement the FL infrastructure across two institutions was assessed by setting collaboration agreements, passing the IT security requirements in both institutions independently, and being able to establish secure connection with both servers at once through R and DataSHIELD. Second, the replication of previous validation efforts was set as a goal for testing the feasibility of federated data harmonization and model training.

## RESULTS

### Study Population

No statistical differences in the distribution of variables were found between the studies for GCS motor scores, pupillary reactivity, hypoxia, Marshall CT scores, traumatic subarachnoid hemorrhage (tSAH), epidural hematoma (EDH), and glucose **(Table 2 and eTable 1)**. Compared with CENTER-TBI, subjects in TRACK-TBI were younger (median [Interquartile range (IQR)]: 37 [25-54] vs. 49 [29-66], p < 0.001), with a lower proportion of subjects with hypotension (54/441, 12% vs. 188/1175, 16%, p = 0.024), and higher hemoglobin levels (median [25-75]: 13.8 [12.5-15.6]) vs. 13 [11.3-14.2], p < 0.001). The proportion of deceased subjects at 6 months was significantly lower in the TRACK-TBI dataset (86/441, 20% vs. 347/1175, 30%, p < 0.001), while the difference in the proportion of unfavorable outcomes at 6 months was similar (216/441, 49% vs 635/1175, 54%, p = 0.078). Compared with the IMPACT development cohort^3^ **(Table 2)**, the combined cohort used for FL was older, with a higher proportion of GCS motor score of 1 or “none” (720/1616, 45% vs. 1395/8509, 16%), a higher proportion of subjects with both pupils reacting (1098/1616, 68% vs. 4486/8509 53%), a higher proportion of Marshall CT classification of II (574/1616, 36% vs. 1838/8509, 22%) and V/VI (543/1616, 34% vs. 1944/8509 23%), and a higher proportion of tSAH (1034/1616, 64% vs. 3313/8509, 39%).

**Table 2.** Summary statistics of each study dataset and the pooled dataset calculated through federated analysis.

| Characteristics | Measure or Category | Development |  |  |  |
| --- | --- | --- | --- | --- | --- |
|  |  | IMPACT*<br>(n = 8509) | TRACK-TBI<br>(n = 441) | CENTER-TBI<br>(n = 1175) | FL-pooled<br>(n = 1616) |
| 6-month outcome | Dead | 2396 (28%) | 86 (20%) | 347 (30%) | 433 (27%) |
|  | Unfavorable | 4082 (48%) | 216 (49%) | 635 (54%) | 851 (53%) |
| Age (years) | Median (IQR) | 30 (21-45) | 37 (25-54) | 49 (29-66) | 45.7 (27.9-62.7) |
| GCS motor score | None (1) | 1395 (16%) | 191 (43%) | 529 (45%) | 720 (45%) |
|  | Extension (2) | 1042 (12%) | 26 (6%) | 65 (6%) | 91 (6%) |
|  | Abnormal flexion (3) | 1085 (13%) | 17 (4%) | 66 (6%) | 83 (5%) |
|  | Normal flexion (4) | 1940 (23%) | 65 (15%) | 118 (10%) | 183 (11%) |
|  | Localizes/obeys (5/6) | 2591 (30%) | 138 (31%) | 397 (34%) | 535 (34%) |
|  | Unknown | 456 (5%) | 4 (1%) | 0 (0%) | 4 (0%) |
| Pupillary reactivity | Both | 4486 (53%) | 279 (63%) | 819 (70%) | 1098 (68%) |
|  | None | 1754 (21%) | 88 (20%) | 216 (18%) | 304 (19%) |
|  | One | 886 (10%) | 29 (7%) | 99 (8%) | 128 (8%) |
|  | Unknown | 1383 (16%) | 45 (10%) | 41 (4%) | 86 (5%) |
| Hypoxia | Yes or suspected | 1116 (13%) | 67 (15%) | 199 (17%) | 266 (17%) |
|  | Unknown | 3057 (36%) | 0 (0%) | 73 (6%) | 73 (5%) |
| Hypotension | Yes or suspected | 1171 (14%) | 54 (12%) | 188 (16%) | 242 (15%) |
|  | Unknown | 2089 (25%) | 0 (0%) | 71 (6%) | 71 (4%) |
| CT classification (Marshall) | I | 360 (4%) | 30 (7%) | 64 (5%) | 94 (6%) |
|  | II | 1838 (22%) | 177 (40%) | 397 (34%) | 574 (36%) |
|  | III/IV | 1050 (12%) | 41 (9%) | 116 (10%) | 157 (10%) |
|  | V/VI | 1944 (23%) | 158 (34%) | 385 (33%) | 543 (34%) |
|  | Unknown | 3317 (39%) | 35 (8%) | 213 (18%) | 248 (15%) |
| tSAH | Yes | 3313 (39%) | 315 (71%) | 719 (61%) | 1034 (64%) |
|  | Unknown | 1116 (13%) | 35 (8%) | 267 (23%) | 302 (19%) |
| EDH | Yes | 999 (12%) | 67 (15%) | 164 (14%) | 231 (14%) |
|  | Unknown | 1100 (13%) | 35 (8%) | 267 (23%) | 302 (19%) |
| Glucose (mmol/l) | Median (25-75) | 8.2 (6.7-10.4) | 7.8 (6.6-9.7) | 7.8 (6.5-9.7) | 7.8 (6.5-9.7) |
|  | Unknown | 3679 (43%) | 26 (6%) | 211 (18%) | 237 (15%) |
| Hemoglobin (g/dl) | Median (25-75) | 12.7 (10.8-14.3) | 13.8 (12.5-15.6) | 13.0 (11.3-14.2) | 13.2 (11.6-14.4) |
|  | Unknown | 4133 (49%) | 18 (4%) | 116 (10%) | 134 (8%) |
| Categorical variables and Unknown: count (%). tSAH: traumatic subarachnoid hemorrhage. EDH: epidural hematoma. *for the IMPACT development dataset, information was extracted and adapted from previous publication<sup>2</sup>. Specifically, % were recalculated respect to full cohort rather than total observed as in the original report for consistency with this work. |  |  |  |  |  |

### Validation of the Original IMPACT TBI Models

When validated on the CENTER-TBI and TRACK-TBI datasets over the federated network, the original IMPACT models presented average AUC values ranging from 0.77 to 0.86 (**Table 3**). The original IMPACT model had the lowest AUC when predicting unfavorable outcomes on the TRACK-TBI dataset using the core predictor sets, and the highest AUC when predicting mortality on the TRACK-TBI dataset using the Extended predictor sets. The calibration performance was variable **(eFigure 1)**, with well-calibrated probabilities for the prediction of unfavorable outcomes on the CENTER-TBI and TRACK-TBI datasets, and miscalibrated probabilities for mortality prediction, especially when predicting the TRACK-TBI dataset. This is also reflected in deviations from ideal calibration intercepts, slopes, and BSs **(eFigure 1 and eTable 2)**.

**Table 3.**
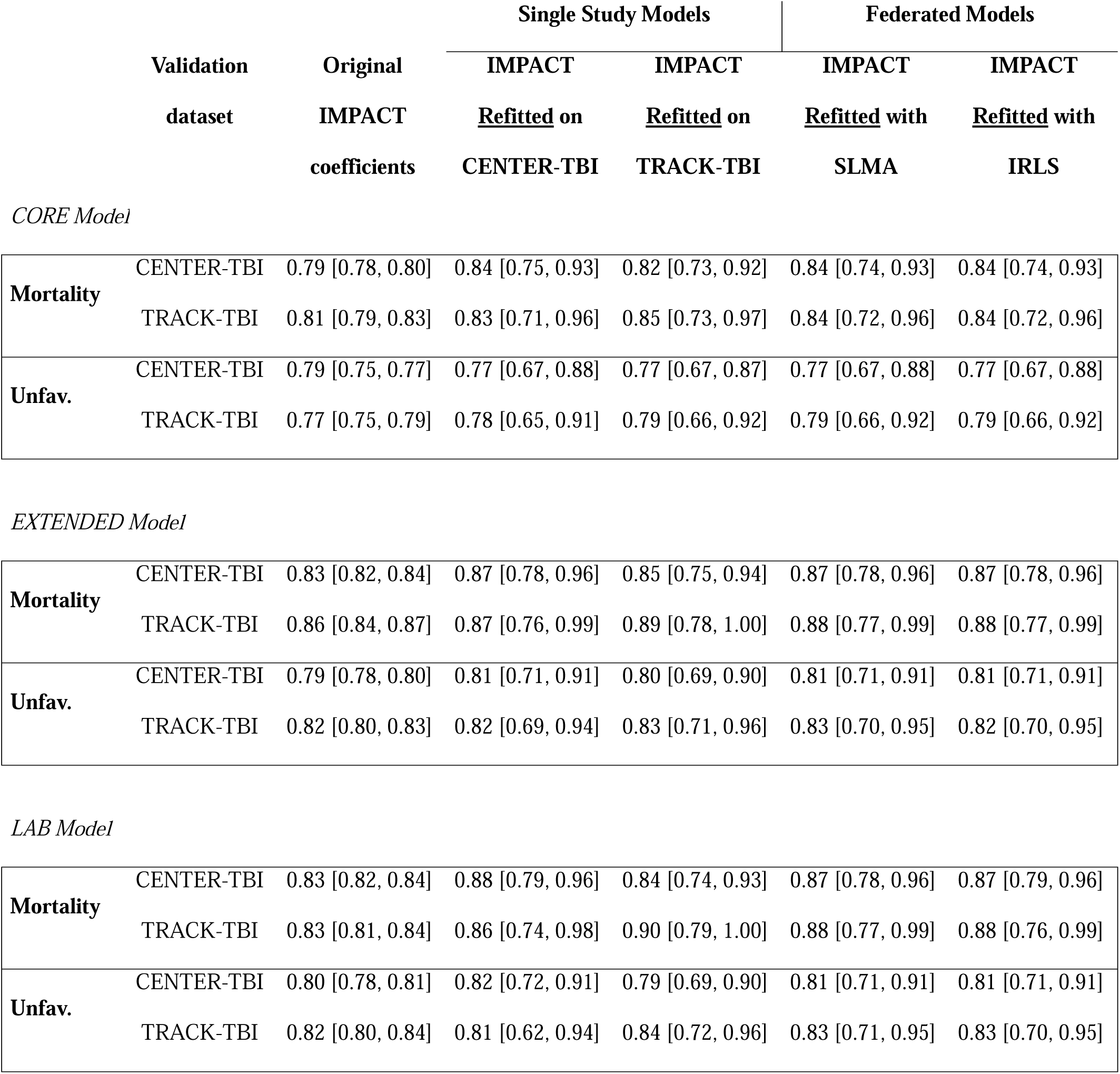
Model discrimination measure (AUC) for all combinations of cross-evaluation. Mean AUC [95%CI] across 5 multiple imputed datasets are presented. Five models are evaluated: Original IMPACT coefficients, the IMPACT model refitted on CENTER-TBI data only, the IMPACT model refitted on TRACK-TBI data only, the IMPACT model refitted on the federated data using SLMA, and the IMPACT model refitted on the federated data using IRLS. Each model approach is validated against the CENTER-TBI or the TRACK-TBI datasets independently. For the Original IMPACT coefficients model, this constitutes an external validation. For the single study models the validation is internal if the dataset for validation is the same than the one used for refitting (eg, refitted in CENTER-TBI and evaluated in CENTER-TBI) and external validation if evaluated on the other study (eg, refitted in TRACK-TBI and evaluated in CENTER-TBI). For both federated models, since the validation is independent for each study, it is considered a subset validation.

### Federated Model Updates

We re-estimated model coefficients to update models over the federated network, resulting in minimal model differences in goodness-of-fit (**eTable 3**) and coefficient estimates **(eFigure 2, eTables 4-15)**. Coefficient standard errors were generally smaller for the FL models than the study-level models **(eFigure 3**, **eTables 4-15)**, indicative of a more precise estimation of model coefficients. Importantly, the effect of missing predictor data on the precision of model estimates was the least in SLMA and IRLS models. (Details on these findings are presented in the Supplementary results.) The AUC values of the updated models ranged between 0.77-0.89, with little difference between the different trained models predicting the same validation dataset **(Table 3)**. As expected, study-level models (CENTER-TBI and TRACK-TBI) performed, in general, slightly better in the training dataset than in the original IMPACT (AUC = 0.77-0.89 vs. 0.77-0.86) and a cross-validated dataset (AUC = 0.77-0.87), although differences were minimal. SLMA and IRLS performed almost identically, predicting the same dataset, with AUC comparable to study-level models and consistently higher than the original IMPACT, except in predicting CENTER-TBI data with the core model for unfavorable outcome.

All models were well-calibrated for predicting CENTER-TBI data **(Figure 2)**. The models with worse calibrations were those trained in the TRACK-TBI data. Some model evaluations predicting TRACK-TBI data show substantial deviations from the ideal calibration, reflected in deviations from the expected calibration intercept and slope, and low BSs scores **(eTable 2)**. More noticeably, the study-level CENTER-TBI model showed deviation from the ideal calibration in predicting TRACK-TBI mortality with the core and laboratory models **(Figure 2 and eTable 2)**. The SLMA and IRLS models generally present well-calibrated predictions **(Figure 2 and eTable 2)**, mitigating some of the miscalibration shown by the CENTER-TBI models when predicting TRACK-TBI outcomes.

## DISCUSSION

In this comprehensive validation and update of the IMPACT prognostic models in 2 large, contemporary multi-continental cohorts of TBI patients over a federated network, we demonstrated feasibility, and report high discriminatory performance but suboptimal calibration. All updated models showed good discriminative power; however, there were suboptimal models in calibrated predicted probabilities when considering study-level cross-evaluation. Both FL approaches for updating coefficients performed similarly well in terms of discrimination and calibration, overcoming some shortcomings of the study-level models. There are a few previous reports using FL in TBI data performed in laboratory-controlled settings.^30–32^ A federated infrastructure adds several benefits to multi-study collaborations, such as privacy-preserving analysis, provenance traceability of analysis, and computational efficiency. To our knowledge, this is the first report on real-world multi-continental implementation of a federated network infrastructure and validation and update of prognostic models in TBI using FL.

The IMPACT models have been extensively externally validated, including in the validation cohorts reported here. We purposefully included the same subjects as in the previous validation studies to benchmark our implementation of a federated infrastructure and FL analysis. Both study-level models produced model coefficients, discrimination, and calibration in line with those reported earlier,^5,6^ indicating that our federated workflow (including data preparation, harmonization, modeling, and evaluation, all performed in a privacy-preserving environment in which the investigator is blinded to implementation of the analysis) is feasible and accurate.

Cross-validation of the study-level models showed differences compared with internal validation. External validation model performance is sensitive to data shift, that is, differences in the cohort characteristics and predictor-outcome associations between the training and the validation cohorts.^33,34^ There are a few substantial differences in predictors’ values (age, hemoglobin, and hypoxia) and outcomes (mortality rates) between the CENTER-TBI and TRACK-TBI cohorts. These may explain the relative reduction in performance and miscalibration when evaluating a study-level model on the other study (external validity).

SLMA and IRLS models were nearly identical in coefficient estimates, discrimination, and calibration performance. Previous studies in pooling IMPACT models across different cohorts have shown similar results, where differences on 1-stage and 2-stage (SLMA) approaches to generate a global model were negligible.^24^ In our analysis, both federated models presented high discrimination performance and calibration similar to the study-level models or higher. The federated models somewhat reduce the high variability in the parameter estimates of some predictors due to low sample representation and missingness. Although expected due to the increase in sample size and meaningful representation during learning, the advantages of a global model across datasets against study-level models should be carefully considered. Pooling model coefficients, as in SLMA, requires similarity between studies to provide good estimates.^24,26^ When between-study heterogeneity due to case-mix regarding observed outcomes and patient characteristics exist, the pooling of estimates into a global model might be biased.^24^ In cases where a well-developed model exists, such as IMPACT, model updates specific to a cohort (or collection of very similar cohorts) might be more reasonable than a global model across distinct cohorts given sufficient sample size.^24^ Where models need updates and a collection of small datasets is available, pooling into a global model might be beneficial if between-study heterogeneity is acceptable.

Although the similarities between SLMA and IRLS suggest that a distributed computing infrastructure might not be necessary since SLMA can be performed by independently fitting models first and aggregating parameter estimates, this conclusion should be considered cautiously.^25^ Aggregated estimates from SLMA are similar to those obtained from pooling individual participant data under conditions where the correlation structure between predictors and between predictors and outcomes is similar.^35^ In that case, training a model across all data sources should produce higher precision estimates because of the gain in sample size. We can observe this in our analysis with the general reduction of coefficient standard errors in the FL models compared to the study-level models. However, if marked between-study heterogeneity exists, coefficients will be biased and not applicable to predict any particular setting.^24^ Since coefficient estimates between IRLS and SLMA were very similar and IRLS generally showed lower standard errors, we must conclude that the observed differences between the TBI cohorts are insufficient to generate substantial deviations between the studied FL approaches. This might not be true in developing or evaluating models using other cohorts or models using the same 2 cohorts, but for predicting different outcomes and/or using different predictor sets. Procedures exist to test before modeling whether pooling will provide an increase in precision in regression estimates with respect to single-study models^35^ and to estimate between-study heterogeneity,^24^ which could be adapted to determine the need for IRLS in the federated context. Therefore, in practice, both co-learning and model pooling federated approaches, such as SLMA, might be required and compared to determine the best approach.

### Limitations and Future Directions

Federated approaches require considerable resources and expertise. Implementing a decentralized network and federated infrastructure is time-consuming and demands tight coordination across sites. Specialized expertise is required for both the programming framework and the study datasets.

Our sub-evaluation might not reflect true generalizability as the federated models have effectively been trained on the totality of the data for comparability with previous validation efforts. Nonetheless, our cross-validation can be considered a particular case of “internal-external cross-validation” among sites.^33^ It is reasonable to expect that federated models trained across multiple datasets are more precise, generalizable, and robust than single-study models when there is little heterogeneity. However, we cannot conclude generalizability. Investigating different methods for pooling estimates in SLMA to update prediction models in TBI may be fruitful.^25^ For instance, the prognostic weights of past learning as captured in a linear predictor could be used as a reference in updating procedures.^36^ Providing cohort descriptions, as well as coefficients, standard error, and covariance matrix estimates openly, could facilitate and accelerate prognostic model updating without the need to share data. While we study model update through the federated network by re-estimating the coefficients, we note that other approaches such as re-calibration could be considered.^4^

## Conclusions

Federated approaches are a feasible alternative to data sharing where global regulatory constraints would hinder collaborations. A decentralized computing network allows for reproducible analytical workflows, FL, cross-validation, and handling multiple imputations for missing values, only subject to limitations on implemented computational capabilities. This initial analysis is our first project to evaluate the federated system, and the return on investment should be valued in the medium-to long-term. Our empirical demonstration of FL’s feasibility, reliability, and utility in TBI across continents opens opportunities for international collaborations to develop federated analysis, such as a multi-study evaluation of the newly proposed CBI-M (clinical, biomarker, imaging, modifier) classification framework,^37^ or more complex prediction models using machine learning and artificial intelligence in TBI.^38–42^

### Transparency, Rigor and Reproducibility Summary

The code reproducing this research is available at XXX (TBD upon acceptance). Data for CENTER-TBI and TRACK-TBI are available under a collaboration request to the respective team coordinators. The study websites provide information on requesting access to data.

## Supporting information

Supplementary meterial

## Author Contributions

A.T.E.: Conceptualization and design; Acquisition, analysis, or interpretation of data; Writing-Original draft, Critical review of the manuscript for important intellectual content; Statistical analysis; Administrative, technical, or material support; Supervision. P.J.B.: Conceptualization and design; Acquisition, analysis, or interpretation of data; Critical review of the manuscript for important intellectual content; Administrative, technical, or material support, Supervision. A.I.R.M.: Conceptualization and design, Acquisition, analysis, or interpretation of data; Critical review of the manuscript for important intellectual content; Funding acquisition; Supervision. G.T.M.: Conceptualization and design; Acquisition, analysis, or interpretation of data; Critical review of the manuscript for important intellectual content; Funding acquisition; Supervision. A.R.F.: Conceptualization and design; Acquisition, analysis, or interpretation of data; Critical review of the manuscript for important intellectual content. J.C.W., T.B.K. and B.P.T.H.: Administrative, technical, or material support; Acquisition, analysis, or interpretation of data; Critical review of the manuscript for important intellectual content. H.E.H, S.J., X.S., J.K.Y., D.P., A.M., H.F.L., A.J.M., D.K.M., E.W.S., CENTER-TBI and TRACK-TBI investigators: Acquisition, analysis, or interpretation of data; Critical review of the manuscript for important intellectual content.

## Consent to participate

Each participant (or their legally authorized representative) provided written informed consent.

## Consent for publication

Not applicable

## Data Availability

Data for CENTER-TBI and TRACK-TBI are available under a collaboration request to the respective team coordinators. The study websites provide information on requesting access to data.

https://tracktbi.ucsf.edu/

https://www.center-tbi.eu/

