## Supplementary meterial for "Use of Federated Learning for validating and updating privacy-preserving decentralized multi-study prognostic models in Traumatic Brain Injury"

**Supplementary material**

**Extra results on comparing models**

eTables 4 to 15 show all the fitted models' coefficient estimates, odds ratios (OR), and measures of missing impact. The relative efficiency (RE), a measure of precision in the parameter estimates due to missingness, ranged between 0.99 and 0.82 and was generally lower in those predictors with a higher proportion of missing data. The proportion of total variance due to missingness (FMI) was relatively low for all the core models, ranging from 0.001 to 0.15. Pupils None and Pupils One had the highest coefficients for the TRACK-TBI and federated models. Notably, the FMI for Pupils One reduces from 0.13 in the TRACK-TBI core model predicting mortality to 0.04 for the same IRLS model. For the extended models, FMI ranged from 0.006 to 0.45, where tSAH has the higher values for all models except for the TRACK-TBI ones. For the lab models, FMI ranged from 0.008 to 0.84. Glucose had the highest FMI for the CENTER-TBI lab model predicting mortality (0.84), followed by tSAH (0.44). On the contrary, the FMI for Glucose in the SLMA lab model predicting mortality was 0.33.

Of the three model sets (core, extended, and laboratory), the only statistical difference in coefficient estimates between models was found for Age between the study level CENTER-TBI and TRACK-TBI laboratory models predicting unfavorable outcomes (z = 1.97, p = 0.048) (eFigure 2). Nonetheless, some important differences were found in the magnitude of the difference between coefficients. Most remarkably, the pooled coefficients for a Marshall CT classification of I were -7.91 and -7.88 for the TRACK-TBI extended and laboratory models predicting mortality, respectively. For the same models, the CENTER-TBI coefficients were -0.19 (extended) and -0.21 (laboratory), the SLMA coefficients were -0.12 (extended) and -0.14 (laboratory), and the IRLS coefficients were -0.04 (extended) and -0.06 (laboratory). These values are consistent with what was reported earlier for TRACK-TBI^26^ and CENTER-TBI^4^ validations. The extreme values in TRACK-TBI are because no cases with a Marshall I for those with GOSE = 1 at 6 months (mortality) are observed in the imputed datasets, causing estimation issues. These extreme differences in Marshall I coefficients were not observed for the extended and laboratory models predicting unfavorable outcomes.

**Supplementary Figures and Tables**

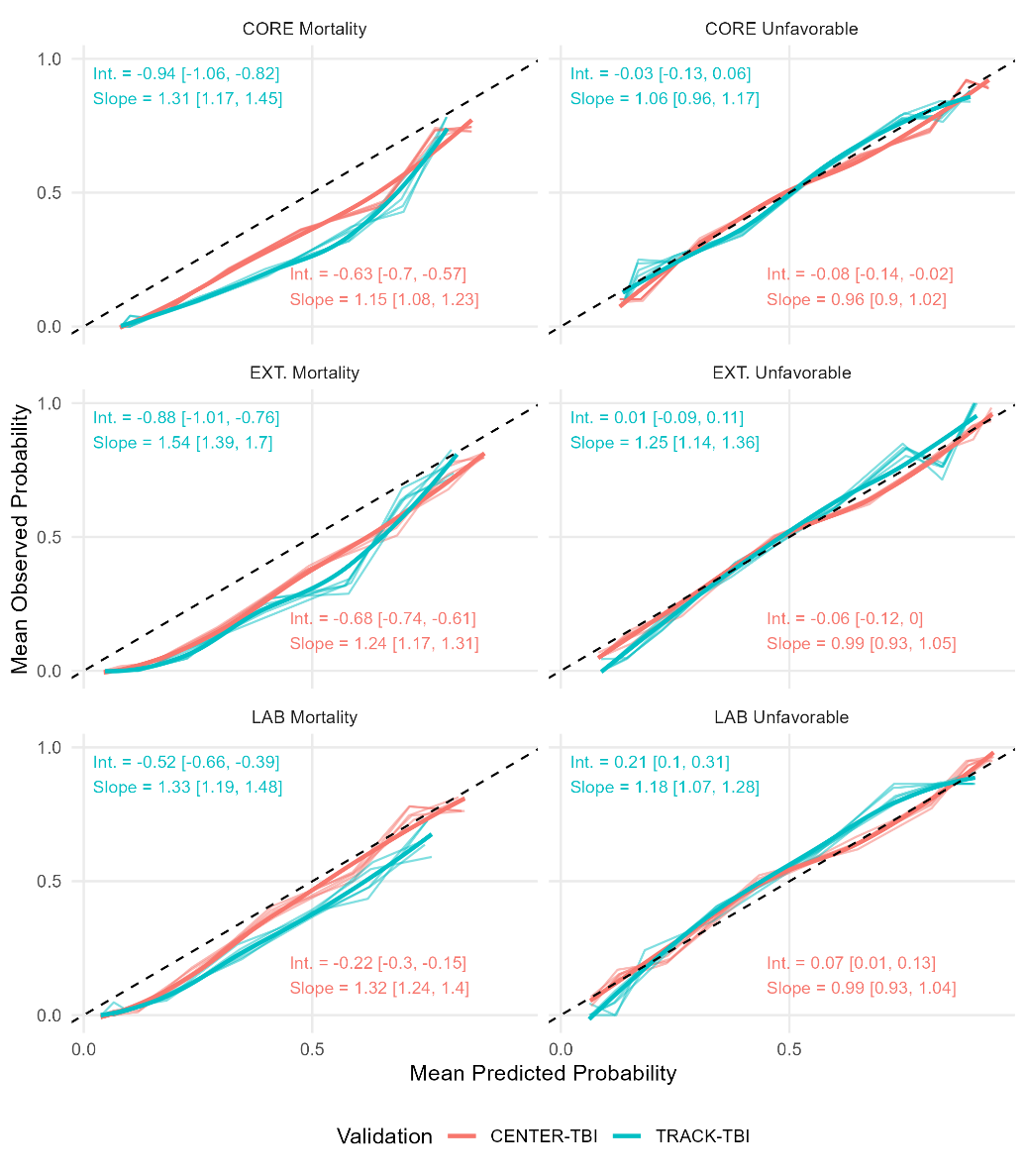

**eFigure 1.** Calibration curves, intercept, and slope for the validation of the original IMPACT coefficients on the CENTER-TBI and TRACK-TBI through the federated infrastructure.

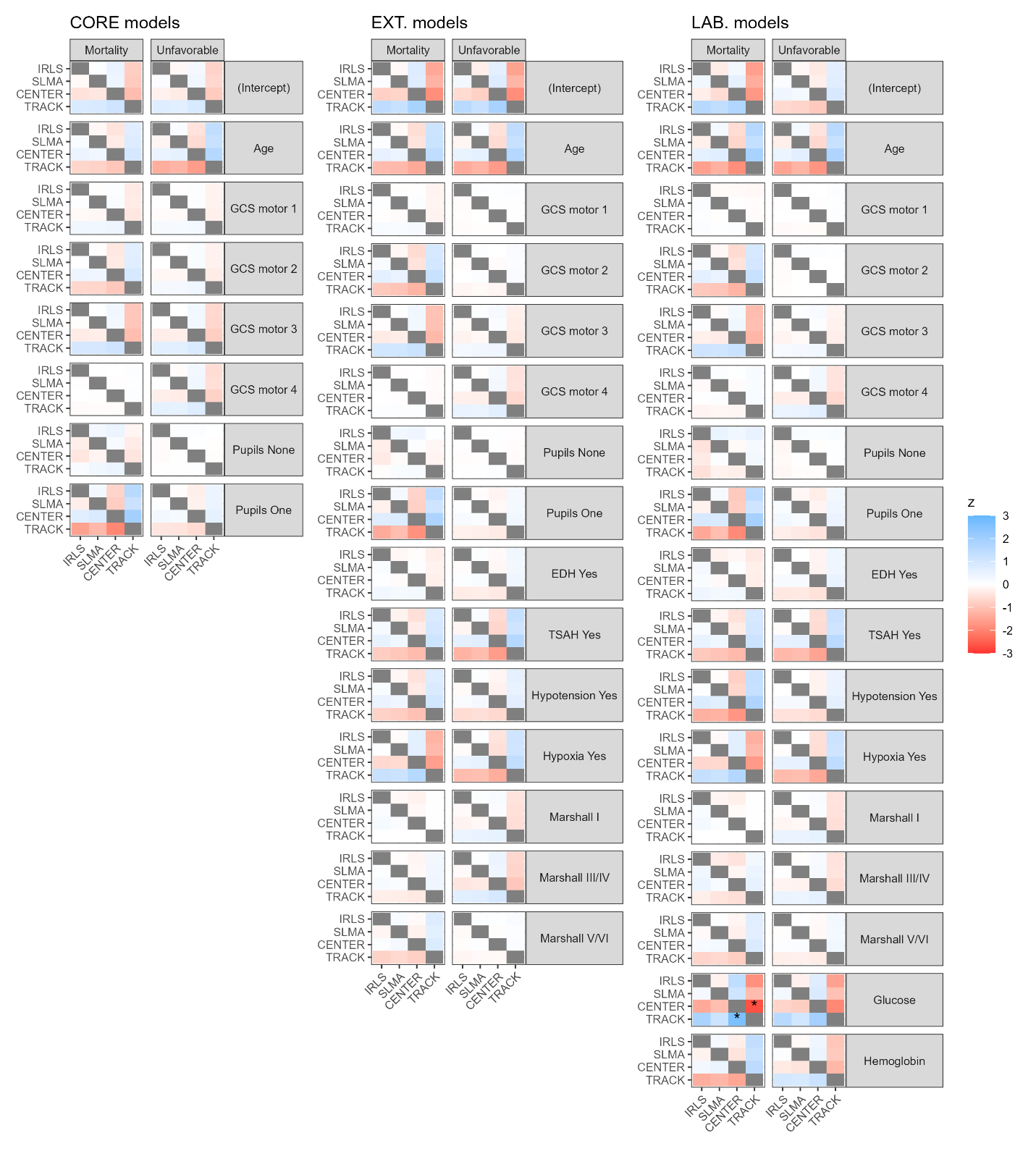

**eFigure 2.** Heatmap of the coefficient standardized mean difference in pairwise comparison between models. *p<0.05

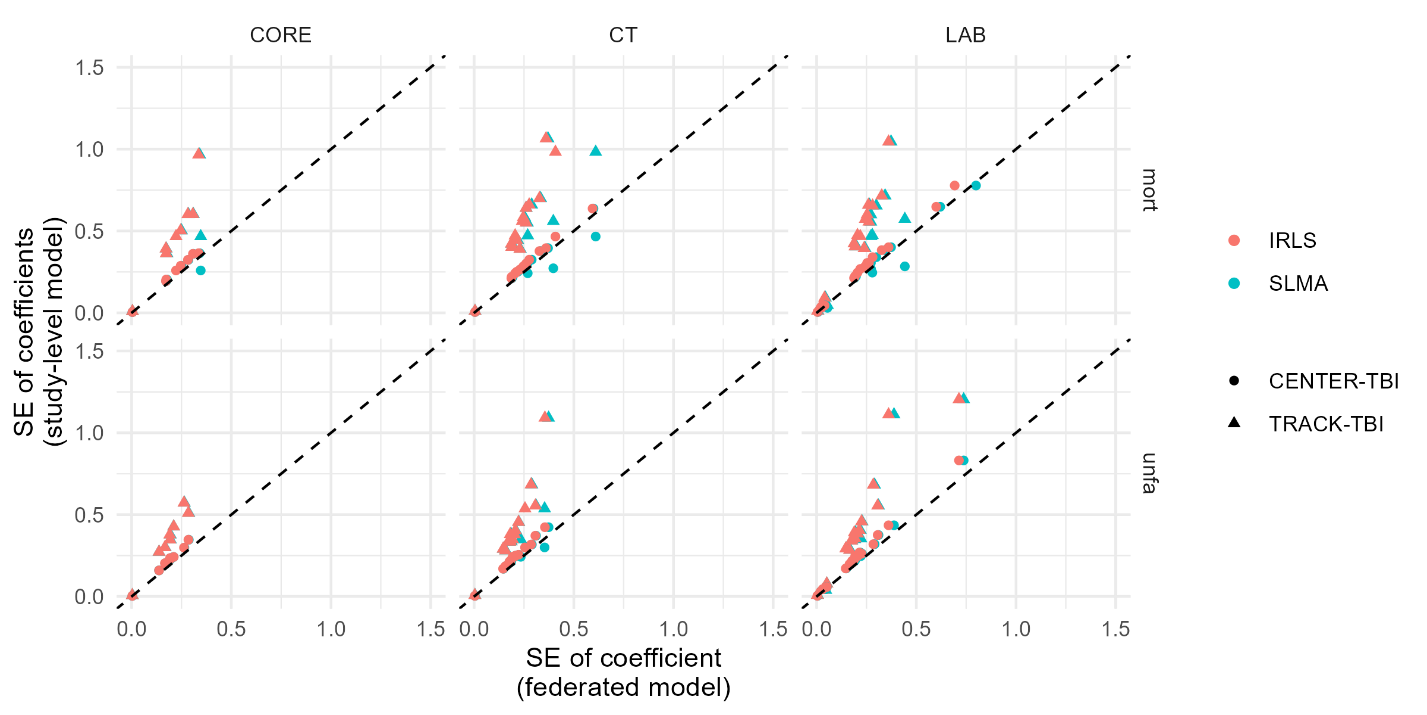

**eFigure 3.** Comparison of the coefficient standard error (SE) for each predictor between the federated models (x-axis) and the study-level models (y-axis) for each one of the six IMPACT model specifications. Alignment around the perfect agreement line (dashed line) would be expected when the SE estimates between the federated models and study-level models are equivalent for each predictor. As observed, the SEs for the study-level models are systematically higher than for the federated models.

**eTable 1.** Summary statistics of each study dataset and P value comparing their differences.

| **Characteristics** | **Measure or Category** | **TRACK-TBI**  **(n = 441)** | **CENTER-TBI**  **(n = 1175)** | **P value** |
| --- | --- | --- | --- | --- |
| Six-month outcome | Dead | 86 (20%) | 347 (30%) | < .001*** |
|  | Unfavorable | 216 (49%) | 635 (54%) | .078 |
| Age (years) | Median (IQR) | 37 (25-54) | 49 (29-66) | < .001*** |
| GCS motor score | None (1) | 191 (43%) | 529 (45%) | .059 |
|  | Extension (2) | 26 (6%) | 65 (6%) |  |
|  | Abnormal flexion (3) | 17 (4%) | 66 (6%) |  |
|  | Normal flexion (4) | 65 (15%) | 118 (10%) |  |
|  | Localizes/obeys (5/6) | 138 (31%) | 397 (34%) |  |
|  | Unknown | 4 (1%) | 0 (0%) |  |
| Pupillary reactivity | Both | 279 (63%) | 819 (70%) | .315 |
|  | None | 88 (20%) | 216 (18%) |  |
|  | One | 29 (7%) | 99 (8%) |  |
|  | Unknown | 45 (10%) | 41 (4%) |  |
| Hypoxia | Yes or suspected | 67 (15%) | 199 (17%) | .203 |
|  | Unknown | 0 (0%) | 73 (6%) |  |
| Hypotension | Yes or suspected | 54 (12%) | 188 (16%) | .024* |
|  | Unknown | 0 (0%) | 71 (6%) |  |
| CT classification  (Marshall) | I | 30 (7%) | 64 (5%) | .651 |
|  | II | 177 (40%) | 397 (34%) |  |
|  | III/IV | 41 (9%) | 116 (10%) |  |
|  | V/VI | 158 (34%) | 385 (33%) |  |
|  | Unknown | 35 (8%) | 213 (18%) |  |
| tSAH | Yes | 315 (71%) | 719 (61%) | .561 |
|  | Unknown | 35 (8%) | 267 (23%) |  |
| EDH | Yes | 67 (15%) | 164 (14%) | .543 |
|  | Unknown | 35 (8%) | 267 (23%) |  |
| Glucose (mmol/l) | Median (IQR) | 7.8 (6.6-9.7) | 7.8 (6.5-9.7) | .112 |
|  | Unknown | 26 (6%) | 211 (18%) |  |
| Hemoglobin (g/dl) | Median (IQR) | 13.8 (12.5-15.6) | 13.0 (11.3-14.2) | < .001*** |
|  | Unknown | 18 (4%) | 116 (10%) |  |
| Categorical variables: count (%), p-value from Fisher’s test. Continuous variables: median (25-75 quartiles), p-value from t-test. Unknown: count (%). tSAH: traumatic subarachnoid hemorrhage. EDH: epidural hematoma | | | | |

**eTable 2.** Scaled Brier Score for predicting the validation datasets with the original IMPACT and refitted models. Mean sBS [95%CI] across five multiple imputed datasets are presented. Five models are evaluated: Original IMPACT coefficients, the IMPACT model refitted on CENTER-TBI data only, the IMPACT model refitted on TRACK-TBI data only, the IMPACT model refitted on the federated data using SLMA, and the IMPACT model refitted on the federated data using IRLS. Each model approach is validated against the CENTER-TBI or the TRACK-TBI datasets independently. For the Original IMPACT coefficients model, this constitutes an external validation. For the single study models the validation is internal if the dataset for validation is the same than the one used for refitting (e.g., refitted in CENTER-TBI and evaluated in CENTER-TBI) and external validation if evaluated on the other study (e.g., refitted in TRACK-TBI and evaluated in CENTER-TBI). For both federated models, since the validation is independent for each study, it is considered a subset validation.

|  |  |  | **Single Study Models** | | **Federated Models** | |
| --- | --- | --- | --- | --- | --- | --- |
|  | **Validation dataset** | **Original IMPACT coefficients** | **IMPACT Refitted on CENTER-TBI** | **IMPACT Refitted on TRACK-TBI** | **IMPACT Refitted with SLMA** | **IMPACT Refitted with IRLS** |
| *CORE Model* | |  |  |  |  |  |
| **Mortality** | CENTER-TBI | 0.15 [0.15-0.15] | 0.31 [0.3-0.31] | 0.27 [0.26-0.27] | 0.3 [0.3-0.3] | 0.3 [0.3-0.3] |
|  | TRACK-TBI | 0.04 [0.03-0.05] | 0.24 [0.23-0.25] | 0.28 [0.28-0.29] | 0.26 [0.25-0.27] | 0.26 [0.26-0.27] |
| **Unfav.** | CENTER-TBI | 0.2 [0.2-0.21] | 0.22 [0.22-0.23] | 0.21 [0.21-0.21] | 0.22 [0.22-0.22] | 0.22 [0.22-0.22] |
|  | TRACK-TBI | 0.22 [0.21-0.22] | 0.24 [0.24-0.25] | 0.25 [0.25-0.26] | 0.25 [0.24-0.25] | 0.24 [0.24-0.25] |
| *EXTENDED Model* | |  |  |  |  |  |
| **Mortality** | CENTER-TBI | 0.21 [0.21-0.22] | 0.38 [0.37-0.38] | 0.31 [0.30-0.32] | 0.36 [0.35-0.38] | 0.37 [0.36-0.38] |
|  | TRACK-TBI | 0.14 [0.13-0.13] | 0.31 [0.31-0.32] | 0.37 [0.37-0.38] | 0.34 [0.33-0.35] | 0.34 [0.34-0.35] |
| **Unfav.** | CENTER-TBI | 0.25 [0.25-0.26] | 0.28 [0.27-0.29] | 0.24 [0.23-0.25] | 0.28 [0.27-0.29] | 0.28 [0.27-0.29] |
|  | TRACK-TBI | 0.3 [0.29-0.3] | 0.3 [0.29-0.31] | 0.33 [0.33-0.34] | 0.32 [0.31-0.32] | 0.31 [0.31-0.32] |
| *LAB Model* |  |  |  |  |  |  |
| **Mortality** | CENTER-TBI | 0.27 [0.26-0.28] | 0.39 [0.38-0.39] | 0.30 [0.28-0.31] | 0.36 [0.34-0.38] | 0.38 [0.37-0.39] |
|  | TRACK-TBI | 0.16 [0.15-0.27] | 0.27 [0.26-0.28] | 0.39 [0.38-0.39] | 0.33 [0.32-0.34] | 0.33 [0.33-0.34] |
| **Unfav.** | CENTER-TBI | 0.26 [0.25-0.27] | 0.3 [0.29-0.3] | 0.23 [0.22-0.24] | 0.28 [0.27-0.3] | 0.29 [0.28-0.3] |
|  | TRACK-TBI | 0.29 [0.29-0.3] | 0.29 [0.28-0.3] | 0.35 [0.34-0.35] | 0.30 [0.29-0.32] | 0.32 [0.31-0.32] |

**eTable 3.** Nagelkerke pseudo-R^2^ for all refitted models. Mean pseudo-R^2^ [95%CI] across five multiple imputed datasets is presented.

|  | **Single Study Models** | | **Federated Models** | |
| --- | --- | --- | --- | --- |
| **Model** | **IMPACT Refitted on CENTER-TBI** | **IMPACT Refitted on TRACK-TBI** | **IMPACT Refitted with SLMA** | **IMPACT Refitted with IRLS** |
| CORE Mortality | 0.4 [0.4, 0.4] | 0.39 [0.38, 0.4] | 0.4 [0.39, 0.4] | 0.39 [0.38, 0.39] |
| CORE Unfavorable | 0.3 [0.3, 0.3] | 0.32 [0.32, 0.33] | 0.3 [0.3, 0.3] | 0.3 [0.3, 0.3] |
| EXT. Mortality | 0.48 [0.47, 0.48] | 0.5 [0.49, 0.5] | 0.48 [0.47, 0.48] | 0.47 [0.46, 0.47] |
| EXT. Unfavorable | 0.36 [0.36, 0.37] | 0.43 [0.42, 0.43] | 0.38 [0.38, 0.39] | 0.37 [0.37, 0.38] |
| LAB Mortality | 0.49 [0.48, 0.5] | 0.5 [0.5, 0.51] | 0.49 [0.49, 0.5] | 0.47 [0.47, 0.48] |
| LAB Unfavorable | 0.38 [0.37, 0.39] | 0.44 [0.43, 0.45] | 0.4 [0.39, 0.41] | 0.38 [0.38, 0.39] |

**eTable 4.** CORE Mortality Study-level model parameters. OR = Odds Ratio, *β =* Model coefficient, SE = Standard Error, FMI = Fraction of missing information, RE = relative efficiency due to missingness, 95% CI = 95% confidence interval of the coefficient

| **TRACK-TBI CORE Mortality** | | | | | | | **CENTER-TBI CORE Mortality** | | | | | |
| --- | --- | --- | --- | --- | --- | --- | --- | --- | --- | --- | --- | --- |
| Term | OR | *β* | *SE* | FMI | RE | 95% CI | OR | *β* | *SE* | FMI | RE | 95% CI |
| Age | 1.07 | 0.07 | 0.01 | 0.06 | 0.99 | [0.05, 0.08] | 1.06 | 0.06 | 0 | 0 | 1 | [0.05, 0.07] |
| GCS motor 1 | 2.02 | 0.7 | 0.39 | 0.03 | 0.99 | [-0.07, 1.47] | 2.4 | 0.87 | 0.19 | 0.01 | 1 | [0.49, 1.25] |
| GCS motor 2 | 3.73 | 1.32 | 0.6 | 0.02 | 1 | [0.13, 2.50] | 1.97 | 0.68 | 0.36 | 0.01 | 1 | [-0.03, 1.38] |
| GCS motor 3 | 0.74 | -0.31 | 0.97 | 0.01 | 1 | [-2.21, 1.59] | 2.24 | 0.8 | 0.36 | 0.01 | 1 | [0.09, 1.52] |
| GCS motor 4 | 1.49 | 0.4 | 0.5 | 0.01 | 1 | [-0.59, 1.39] | 1.43 | 0.36 | 0.29 | 0.02 | 1 | [-0.21, 0.92] |
| Pupils One | 6.36 | 1.85 | 0.47 | 0.06 | 0.99 | [0.93, 2.77] | 2.24 | 0.8 | 0.26 | 0.05 | 0.99 | [0.30, 1.31] |
| Pupils None | 6.9 | 1.93 | 0.36 | 0.13 | 0.97 | [1.21, 2.65] | 8.27 | 2.11 | 0.2 | 0.01 | 1 | [1.71, 2.51] |

**eTable 5.** CORE Mortality federated data model parameters. OR = Odds Ratio, *β =* Model coefficient, SE = Standard Error, FMI = Fraction of missing information, RE = relative efficiency due to missingness, 95% CI = 95% confidence interval of the coefficient

| **SLMA CORE Mortality** | | | | | | | **IRLS CORE Mortality** | | | | | |
| --- | --- | --- | --- | --- | --- | --- | --- | --- | --- | --- | --- | --- |
| Term | OR | *β* | *SE* | FMI | RE | 95% CI | OR | *β* | *SE* | FMI | RE | 95% CI |
| Age | 1.06 | 0.06 | 0 | 0.02 | 1 | [0.05, 0.07] | 1.06 | 0.06 | 0 | 0.01 | 1 | [0.05, 0.07] |
| GCS motor 1 | 2.32 | 0.84 | 0.17 | 0.01 | 1 | [0.50, 1.18] | 2.34 | 0.85 | 0.17 | 0 | 1 | [0.51, 1.19] |
| GCS motor 2 | 2.33 | 0.85 | 0.31 | 0.01 | 1 | [0.24, 1.45] | 2.37 | 0.86 | 0.31 | 0.01 | 1 | [0.26, 1.47] |
| GCS motor 3 | 1.95 | 0.67 | 0.34 | 0.01 | 1 | [-0.00, 1.34] | 1.91 | 0.65 | 0.34 | 0.02 | 1 | [-0.01, 1.31] |
| GCS motor 4 | 1.44 | 0.37 | 0.25 | 0.02 | 1 | [-0.12, 0.86] | 1.43 | 0.36 | 0.25 | 0.01 | 1 | [-0.12, 0.84] |
| Pupils One | 3.2 | 1.16 | 0.35 | 0.06 | 0.99 | [0.48, 1.85] | 2.85 | 1.05 | 0.22 | 0.02 | 1 | [0.61, 1.48] |
| Pupils None | 7.88 | 2.06 | 0.18 | 0.04 | 0.99 | [1.72, 2.41] | 7.37 | 2 | 0.17 | 0.04 | 0.99 | [1.66, 2.34] |

**eTable 6.** CORE Unfavorable outcome Study-level model parameters. OR = Odds Ratio, *β =* Model coefficient, SE = Standard Error, FMI = Fraction of missing information, RE = relative efficiency due to missingness, 95% CI = 95% confidence interval of the coefficient

| **TRACK-TBI CORE Unfavorable** | | | | | | | **CENTER-TBI CORE Unfavorable** | | | | | |
| --- | --- | --- | --- | --- | --- | --- | --- | --- | --- | --- | --- | --- |
| Term | OR | *β* | *SE* | FMI | RE | 95% CI | OR | *β* | *SE* | FMI | RE | 95% CI |
| Age | 1.06 | 0.05 | 0.01 | 0.01 | 1 | [0.04, 0.07] | 1.04 | 0.04 | 0 | 0 | 1 | [0.03, 0.05] |
| GCS motor 1 | 2.5 | 0.92 | 0.27 | 0.02 | 1 | [0.38, 1.45] | 2.71 | 1 | 0.16 | 0 | 1 | [0.69, 1.31] |
| GCS motor 2 | 3.94 | 1.37 | 0.51 | 0.02 | 1 | [0.37, 2.37] | 4.81 | 1.57 | 0.35 | 0 | 1 | [0.89, 2.25] |
| GCS motor 3 | 1.62 | 0.48 | 0.57 | 0.01 | 1 | [-0.64, 1.61] | 2.76 | 1.01 | 0.3 | 0 | 1 | [0.43, 1.60] |
| GCS motor 4 | 1.28 | 0.25 | 0.35 | 0.01 | 1 | [-0.43, 0.93] | 1.76 | 0.56 | 0.23 | 0.01 | 1 | [0.10, 1.02] |
| Pupils One | 2.88 | 1.06 | 0.43 | 0.03 | 0.99 | [0.22, 1.90] | 2.15 | 0.77 | 0.24 | 0.01 | 1 | [0.29, 1.24] |
| Pupils None | 3.91 | 1.36 | 0.3 | 0.09 | 0.98 | [0.77, 1.96] | 3.96 | 1.38 | 0.2 | 0.01 | 1 | [0.98, 1.78] |

**eTable 7.** CORE Unfavorable federated data model parameters. OR = Odds Ratio, *β =* Model coefficient, SE = Standard Error, FMI = Fraction of missing information, RE = relative efficiency due to missingness, 95% CI = 95% confidence interval of the coefficient

| **SLMA CORE Unfavorable** | | | | | | | **IRLS CORE Unfavorable** | | | | | |
| --- | --- | --- | --- | --- | --- | --- | --- | --- | --- | --- | --- | --- |
| Term | OR | *β* | *SE* | FMI | RE | 95% CI | OR | *β* | *SE* | FMI | RE | 95% CI |
| Age | 1.05 | 0.04 | 0 | 0.01 | 1 | [0.04, 0.05] | 1.04 | 0.04 | 0 | 0 | 1 | [0.04, 0.05] |
| GCS motor 1 | 2.66 | 0.98 | 0.14 | 0.01 | 1 | [0.71, 1.25] | 2.68 | 0.98 | 0.14 | 0.01 | 1 | [0.71, 1.25] |
| GCS motor 2 | 4.51 | 1.51 | 0.29 | 0.01 | 1 | [0.94, 2.07] | 4.53 | 1.51 | 0.28 | 0.01 | 1 | [0.95, 2.07] |
| GCS motor 3 | 2.46 | 0.9 | 0.26 | 0.01 | 1 | [0.38, 1.42] | 2.45 | 0.89 | 0.26 | 0.01 | 1 | [0.38, 1.41] |
| GCS motor 4 | 1.59 | 0.46 | 0.19 | 0.01 | 1 | [0.08, 0.85] | 1.62 | 0.48 | 0.19 | 0.01 | 1 | [0.10, 0.86] |
| Pupils One | 2.31 | 0.84 | 0.21 | 0.02 | 1 | [0.42, 1.25] | 2.35 | 0.85 | 0.21 | 0.03 | 0.99 | [0.44, 1.27] |
| Pupils None | 3.95 | 1.37 | 0.17 | 0.04 | 0.99 | [1.04, 1.70] | 3.89 | 1.36 | 0.17 | 0.04 | 0.99 | [1.03, 1.69] |

**eTable 8.** Ext. Mortality Study-level model parameters. OR = Odds Ratio, *β =* Model coefficient, SE = Standard Error, FMI = Fraction of missing information, RE = relative efficiency due to missingness, 95% CI = 95% confidence interval of the coefficient

| **TRACK-TBI EXT. Mortality** | | | | | | | **CENTER-TBI EXT. Mortality** | | | | | |
| --- | --- | --- | --- | --- | --- | --- | --- | --- | --- | --- | --- | --- |
| Term | OR | *β* | *SE* | FMI | RE | 95% CI | OR | *β* | *SE* | FMI | RE | 95% CI |
| Age | 1.07 | 0.07 | 0.01 | 0.07 | 0.99 | [0.05, 0.09] | 1.05 | 0.05 | 0.01 | 0.05 | 0.99 | [0.04, 0.06] |
| GCS motor 1 | 1.78 | 0.58 | 0.42 | 0.05 | 0.99 | [-0.25, 1.41] | 1.96 | 0.67 | 0.21 | 0.03 | 0.99 | [0.26, 1.09] |
| GCS motor 2 | 3.69 | 1.31 | 0.7 | 0.08 | 0.98 | [-0.07, 2.68] | 1.36 | 0.31 | 0.38 | 0 | 1 | [-0.43, 1.05] |
| GCS motor 3 | 0.43 | -0.85 | 1.06 | 0.01 | 1 | [-2.94, 1.25] | 1.62 | 0.48 | 0.4 | 0 | 1 | [-0.29, 1.26] |
| GCS motor 4 | 1.28 | 0.25 | 0.55 | 0.06 | 0.99 | [-0.84, 1.33] | 1.34 | 0.3 | 0.3 | 0.03 | 0.99 | [-0.29, 0.89] |
| EDH Yes | 0.35 | -1.06 | 0.64 | 0.38 | 0.93 | [-2.37, 0.24] | 0.41 | -0.89 | 0.29 | 0.34 | 0.94 | [-1.46, -0.31] |
| Hypotension Yes | 2.21 | 0.79 | 0.44 | 0.02 | 1 | [-0.08, 1.67] | 1.3 | 0.26 | 0.25 | 0.14 | 0.97 | [-0.23, 0.76] |
| Hypoxia Yes | 0.76 | -0.27 | 0.47 | 0.09 | 0.98 | [-1.20, 0.66] | 1.77 | 0.57 | 0.24 | 0.14 | 0.97 | [0.10, 1.05] |
| Marshall I | 0 | -7.7 | 827.24 | 0 | 1 | [-1633.70, 1618.30] | 0.93 | -0.07 | 0.64 | 0.42 | 0.92 | [-1.38, 1.24] |
| Marshall III/IV | 7.46 | 2.01 | 0.58 | 0.09 | 0.98 | [0.86, 3.16] | 5.87 | 1.77 | 0.28 | 0.11 | 0.98 | [1.21, 2.33] |
| Marshall V/VI | 5.54 | 1.71 | 0.39 | 0.1 | 0.98 | [0.94, 2.48] | 3.83 | 1.34 | 0.26 | 0.45 | 0.92 | [0.80, 1.89] |
| Pupils One | 5.74 | 1.75 | 0.56 | 0.22 | 0.96 | [0.63, 2.86] | 1.89 | 0.64 | 0.27 | 0.03 | 0.99 | [0.10, 1.17] |
| Pupils None | 5.64 | 1.73 | 0.4 | 0.14 | 0.97 | [0.93, 2.52] | 6.12 | 1.81 | 0.22 | 0.04 | 0.99 | [1.38, 2.24] |
| TSAH Yes | 2.93 | 1.08 | 0.66 | 0.22 | 0.96 | [-0.24, 2.39] | 1.29 | 0.26 | 0.32 | 0.33 | 0.94 | [-0.40, 0.91] |

**eTable 9.** Ext. Mortality federated data model parameters. OR = Odds Ratio, *β =* Model coefficient, SE = Standard Error, FMI = Fraction of missing information, RE = relative efficiency due to missingness, 95% CI = 95% confidence interval of the coefficient

| **SLMA EXT. Mortality** | | | | | | | **IRLS EXT. Mortality** | | | | | |
| --- | --- | --- | --- | --- | --- | --- | --- | --- | --- | --- | --- | --- |
| Term | OR | *β* | *SE* | FMI | RE | 95% CI | OR | *β* | *SE* | FMI | RE | 95% CI |
| Age | 1.06 | 0.06 | 0 | 0.04 | 0.99 | [0.05, 0.07] | 1.06 | 0.06 | 0 | 0.04 | 0.99 | [0.05, 0.07] |
| GCS motor 1 | 1.92 | 0.65 | 0.19 | 0.03 | 0.99 | [0.28, 1.02] | 1.94 | 0.66 | 0.19 | 0.03 | 0.99 | [0.30, 1.03] |
| GCS motor 2 | 1.73 | 0.55 | 0.33 | 0.03 | 0.99 | [-0.11, 1.20] | 1.85 | 0.61 | 0.33 | 0.02 | 1 | [-0.03, 1.26] |
| GCS motor 3 | 1.38 | 0.32 | 0.37 | 0.01 | 1 | [-0.41, 1.05] | 1.36 | 0.31 | 0.36 | 0 | 1 | [-0.40, 1.01] |
| GCS motor 4 | 1.33 | 0.28 | 0.27 | 0.06 | 0.99 | [-0.24, 0.81] | 1.34 | 0.29 | 0.26 | 0.05 | 0.99 | [-0.22, 0.81] |
| EDH Yes | 0.4 | -0.91 | 0.26 | 0.35 | 0.93 | [-1.44, -0.38] | 0.42 | -0.86 | 0.26 | 0.38 | 0.93 | [-1.39, -0.33] |
| Hypotension Yes | 1.46 | 0.38 | 0.22 | 0.11 | 0.98 | [-0.05, 0.81] | 1.53 | 0.42 | 0.21 | 0.11 | 0.98 | [0.00, 0.84] |
| Hypoxia Yes | 1.42 | 0.35 | 0.27 | 0.11 | 0.98 | [-0.18, 0.88] | 1.45 | 0.37 | 0.21 | 0.1 | 0.98 | [-0.03, 0.78] |
| Marshall I | 1.03 | 0.03 | 0.6 | 0.38 | 0.93 | [-1.19, 1.25] | 1.13 | 0.12 | 0.59 | 0.42 | 0.92 | [-1.10, 1.34] |
| Marshall III/IV | 6.14 | 1.81 | 0.25 | 0.1 | 0.98 | [1.32, 2.31] | 6.26 | 1.83 | 0.25 | 0.09 | 0.98 | [1.35, 2.32] |
| Marshall V/VI | 4.16 | 1.43 | 0.23 | 0.45 | 0.92 | [0.95, 1.91] | 3.93 | 1.37 | 0.23 | 0.47 | 0.91 | [0.89, 1.84] |
| Pupils One | 2.74 | 1.01 | 0.4 | 0.17 | 0.97 | [0.22, 1.79] | 2.47 | 0.9 | 0.24 | 0.05 | 0.99 | [0.43, 1.38] |
| Pupils None | 6 | 1.79 | 0.19 | 0.07 | 0.99 | [1.41, 2.17] | 5.54 | 1.71 | 0.19 | 0.08 | 0.98 | [1.34, 2.08] |
| TSAH Yes | 1.48 | 0.39 | 0.29 | 0.3 | 0.94 | [-0.19, 0.97] | 1.61 | 0.48 | 0.28 | 0.25 | 0.95 | [-0.07, 1.03] |

**eTable 10.** Ext. Unfavorable outcome Study-level model parameters. OR = Odds Ratio, *β =* Model coefficient, SE = Standard Error, FMI = Fraction of missing information, RE = relative efficiency due to missingness, 95% CI = 95% confidence interval of the coefficient

| **TRACK-TBI EXT. Unfavorable** | | | | | | | **CENTER-TBI EXT. Unfavorable** | | | | | |
| --- | --- | --- | --- | --- | --- | --- | --- | --- | --- | --- | --- | --- |
| Term | OR | *β* | *SE* | FMI | RE | 95% CI | OR | *β* | *SE* | FMI | RE | 95% CI |
| Age | 1.05 | 0.05 | 0.01 | 0.05 | 0.99 | [0.03, 0.07] | 1.04 | 0.04 | 0 | 0.02 | 1 | [0.03, 0.04] |
| GCS motor 1 | 2.35 | 0.85 | 0.29 | 0.03 | 0.99 | [0.28, 1.43] | 2.33 | 0.84 | 0.17 | 0.01 | 1 | [0.51, 1.18] |
| GCS motor 2 | 4.51 | 1.51 | 0.55 | 0.08 | 0.98 | [0.41, 2.60] | 4.1 | 1.41 | 0.37 | 0.03 | 0.99 | [0.68, 2.14] |
| GCS motor 3 | 1.82 | 0.6 | 0.68 | 0.06 | 0.99 | [-0.75, 1.94] | 2.4 | 0.88 | 0.32 | 0.01 | 1 | [0.25, 1.50] |
| GCS motor 4 | 1.18 | 0.16 | 0.38 | 0.08 | 0.98 | [-0.58, 0.91] | 1.57 | 0.45 | 0.24 | 0.01 | 1 | [-0.03, 0.93] |
| EDH Yes | 0.52 | -0.66 | 0.33 | 0.12 | 0.98 | [-1.31, 0.00] | 0.45 | -0.8 | 0.21 | 0.23 | 0.96 | [-1.22, -0.38] |
| Hypotension Yes | 2.64 | 0.97 | 0.38 | 0.01 | 1 | [0.22, 1.72] | 1.99 | 0.69 | 0.21 | 0.04 | 0.99 | [0.27, 1.10] |
| Hypoxia Yes | 1.65 | 0.5 | 0.35 | 0.02 | 1 | [-0.18, 1.19] | 0.95 | -0.05 | 0.21 | 0.07 | 0.99 | [-0.46, 0.36] |
| Marshall I | 0.33 | -1.11 | 1.09 | 0.37 | 0.93 | [-3.33, 1.11] | 0.62 | -0.47 | 0.42 | 0.36 | 0.93 | [-1.33, 0.39] |
| Marshall III/IV | 1.79 | 0.58 | 0.4 | 0.03 | 0.99 | [-0.21, 1.38] | 2.79 | 1.03 | 0.25 | 0.11 | 0.98 | [0.53, 1.52] |
| Marshall V/VI | 2.63 | 0.97 | 0.28 | 0.09 | 0.98 | [0.41, 1.52] | 2.53 | 0.93 | 0.19 | 0.26 | 0.95 | [0.56, 1.30] |
| Pupils One | 2.22 | 0.8 | 0.45 | 0.07 | 0.99 | [-0.10, 1.69] | 1.87 | 0.62 | 0.25 | 0.03 | 0.99 | [0.12, 1.12] |
| Pupils None | 2.95 | 1.08 | 0.34 | 0.14 | 0.97 | [0.42, 1.75] | 3.03 | 1.11 | 0.22 | 0.05 | 0.99 | [0.68, 1.54] |
| TSAH Yes | 2.61 | 0.96 | 0.35 | 0.01 | 1 | [0.27, 1.64] | 1.31 | 0.27 | 0.24 | 0.31 | 0.94 | [-0.22, 0.76] |

**eTable 11**. Ext. Unfavorable outcome federated data model parameters. OR = Odds Ratio, *β =* Model coefficient, SE = Standard Error, FMI = Fraction of missing information, RE = relative efficiency due to missingness, 95% CI = 95% confidence interval of the coefficient

| **SLMA EXT. Unfavorable** | | | | | | | **IRLS EXT. Unfavorable** | | | | | |
| --- | --- | --- | --- | --- | --- | --- | --- | --- | --- | --- | --- | --- |
| Term | OR | *β* | *SE* | FMI | RE | 95% CI | OR | *β* | *SE* | FMI | RE | 95% CI |
| Age | 1.04 | 0.04 | 0 | 0.05 | 0.99 | [0.03, 0.05] | 1.04 | 0.04 | 0 | 0.01 | 1 | [0.03, 0.04] |
| GCS motor 1 | 2.33 | 0.85 | 0.15 | 0.02 | 1 | [0.56, 1.14] | 2.3 | 0.83 | 0.15 | 0.02 | 1 | [0.55, 1.12] |
| GCS motor 2 | 4.23 | 1.44 | 0.31 | 0.06 | 0.99 | [0.83, 2.05] | 4.11 | 1.41 | 0.31 | 0.05 | 0.99 | [0.81, 2.02] |
| GCS motor 3 | 2.28 | 0.83 | 0.29 | 0.02 | 1 | [0.26, 1.39] | 2.2 | 0.79 | 0.29 | 0.03 | 0.99 | [0.23, 1.35] |
| GCS motor 4 | 1.44 | 0.36 | 0.21 | 0.04 | 0.99 | [-0.04, 0.77] | 1.45 | 0.37 | 0.2 | 0.04 | 0.99 | [-0.03, 0.77] |
| EDH Yes | 0.47 | -0.76 | 0.18 | 0.19 | 0.96 | [-1.11, -0.41] | 0.46 | -0.78 | 0.18 | 0.19 | 0.96 | [-1.13, -0.43] |
| Hypotension Yes | 2.12 | 0.75 | 0.19 | 0.04 | 0.99 | [0.39, 1.12] | 2.07 | 0.73 | 0.18 | 0.03 | 0.99 | [0.37, 1.08] |
| Hypoxia Yes | 1.09 | 0.09 | 0.18 | 0.06 | 0.99 | [-0.26, 0.44] | 1.08 | 0.07 | 0.18 | 0.07 | 0.99 | [-0.27, 0.42] |
| Marshall I | 0.58 | -0.54 | 0.37 | 0.29 | 0.94 | [-1.29, 0.21] | 0.56 | -0.58 | 0.35 | 0.27 | 0.95 | [-1.29, 0.13] |
| Marshall III/IV | 2.49 | 0.91 | 0.21 | 0.06 | 0.99 | [0.50, 1.32] | 2.43 | 0.89 | 0.21 | 0.07 | 0.99 | [0.48, 1.29] |
| Marshall V/VI | 2.56 | 0.94 | 0.16 | 0.25 | 0.95 | [0.62, 1.25] | 2.51 | 0.92 | 0.16 | 0.26 | 0.95 | [0.61, 1.23] |
| Pupils One | 1.95 | 0.67 | 0.22 | 0.05 | 0.99 | [0.23, 1.11] | 1.98 | 0.68 | 0.22 | 0.06 | 0.99 | [0.25, 1.12] |
| Pupils None | 3 | 1.1 | 0.19 | 0.1 | 0.98 | [0.73, 1.47] | 2.98 | 1.09 | 0.18 | 0.1 | 0.98 | [0.73, 1.45] |
| TSAH Yes | 1.65 | 0.5 | 0.23 | 0.06 | 0.99 | [0.04, 0.96] | 1.58 | 0.46 | 0.19 | 0.19 | 0.96 | [0.07, 0.84] |

**eTable 12.** LAB Mortality Study-level model parameters. OR = Odds Ratio, *β =* Model coefficient, SE = Standard Error, FMI = Fraction of missing information, RE = relative efficiency due to missingness, 95% CI = 95% confidence interval of the coefficient

| **TRACK-TBI LAB Mortality** | | | | | | | **CENTER-TBI LAB Mortality** | | | | | |
| --- | --- | --- | --- | --- | --- | --- | --- | --- | --- | --- | --- | --- |
| Term | OR | *β* | *SE* | FMI | RE | 95% CI | OR | *β* | *SE* | FMI | RE | 95% CI |
| Age | 1.08 | 0.08 | 0.01 | 0.03 | 0.99 | [0.05, 0.10] | 1.05 | 0.05 | 0.01 | 0.07 | 0.99 | [0.04, 0.06] |
| GCS motor 1 | 1.81 | 0.59 | 0.42 | 0.05 | 0.99 | [-0.24, 1.42] | 1.86 | 0.62 | 0.21 | 0.02 | 1 | [0.21, 1.04] |
| GCS motor 2 | 3.85 | 1.35 | 0.72 | 0.08 | 0.98 | [-0.06, 2.76] | 1.39 | 0.33 | 0.38 | 0.01 | 1 | [-0.42, 1.08] |
| GCS motor 3 | 0.4 | -0.92 | 1.05 | 0.01 | 1 | [-2.98, 1.13] | 1.53 | 0.43 | 0.4 | 0.02 | 1 | [-0.36, 1.21] |
| GCS motor 4 | 1.44 | 0.37 | 0.56 | 0.06 | 0.99 | [-0.73, 1.46] | 1.29 | 0.26 | 0.31 | 0.03 | 0.99 | [-0.34, 0.86] |
| EDH Yes | 0.33 | -1.12 | 0.66 | 0.39 | 0.93 | [-2.47, 0.23] | 0.4 | -0.91 | 0.28 | 0.32 | 0.94 | [-1.48, -0.34] |
| Glucose | 0.96 | -0.04 | 0.05 | 0.23 | 0.96 | [-0.13, 0.06] | 1.12 | 0.11 | 0.03 | 0.19 | 0.96 | [0.05, 0.17] |
| Hemoglobin | 1.15 | 0.14 | 0.09 | 0.11 | 0.98 | [-0.04, 0.32] | 0.99 | -0.01 | 0.05 | 0.28 | 0.95 | [-0.10, 0.08] |
| Hypotension Yes | 2.78 | 1.02 | 0.47 | 0.01 | 1 | [0.10, 1.94] | 1.09 | 0.08 | 0.27 | 0.2 | 0.96 | [-0.45, 0.61] |
| Hypoxia Yes | 0.73 | -0.31 | 0.47 | 0.08 | 0.98 | [-1.24, 0.62] | 1.78 | 0.58 | 0.25 | 0.15 | 0.97 | [0.09, 1.06] |
| Marshall I | 0 | -7.84 | 815.87 | 0 | 1 | [-1611.51, 1595.84] | 0.92 | -0.08 | 0.65 | 0.43 | 0.92 | [-1.41, 1.25] |
| Marshall III/IV | 6.93 | 1.94 | 0.6 | 0.1 | 0.98 | [0.75, 3.12] | 4.86 | 1.58 | 0.3 | 0.2 | 0.96 | [0.98, 2.18] |
| Marshall V/VI | 5.35 | 1.68 | 0.39 | 0.11 | 0.98 | [0.90, 2.46] | 3.63 | 1.29 | 0.28 | 0.49 | 0.91 | [0.71, 1.87] |
| Pupils One | 5.85 | 1.77 | 0.57 | 0.23 | 0.96 | [0.62, 2.91] | 1.72 | 0.54 | 0.28 | 0.08 | 0.98 | [-0.01, 1.10] |
| Pupils None | 6.58 | 1.88 | 0.4 | 0.06 | 0.99 | [1.09, 2.68] | 5.81 | 1.76 | 0.22 | 0.04 | 0.99 | [1.32, 2.20] |
| TSAH Yes | 3.01 | 1.1 | 0.65 | 0.21 | 0.96 | [-0.20, 2.40] | 1.28 | 0.25 | 0.34 | 0.38 | 0.93 | [-0.44, 0.94] |

**eTable 13.** LAB Mortality federated data model parameters. OR = Odds Ratio, *β =* Model coefficient, SE = Standard Error, FMI = Fraction of missing information, RE = relative efficiency due to missingness, 95% CI = 95% confidence interval of the coefficient

| **SLMA LAB Mortality** | | | | | | | **IRLS LAB Mortality** | | | | | |
| --- | --- | --- | --- | --- | --- | --- | --- | --- | --- | --- | --- | --- |
| Term | OR | *β* | *SE* | FMI | RE | 95% CI | OR | *β* | *SE* | FMI | RE | 95% CI |
| Age | 1.06 | 0.06 | 0.01 | 0.09 | 0.98 | [0.04, 0.07] | 1.06 | 0.06 | 0 | 0.05 | 0.99 | [0.05, 0.06] |
| GCS motor 1 | 1.85 | 0.62 | 0.19 | 0.03 | 0.99 | [0.24, 0.99] | 1.91 | 0.64 | 0.19 | 0.03 | 0.99 | [0.28, 1.01] |
| GCS motor 2 | 1.78 | 0.58 | 0.34 | 0.02 | 1 | [-0.10, 1.25] | 1.89 | 0.64 | 0.33 | 0.01 | 1 | [-0.00, 1.28] |
| GCS motor 3 | 1.29 | 0.26 | 0.37 | 0.01 | 1 | [-0.48, 0.99] | 1.34 | 0.29 | 0.36 | 0.01 | 1 | [-0.42, 1.00] |
| GCS motor 4 | 1.33 | 0.28 | 0.27 | 0.07 | 0.99 | [-0.25, 0.82] | 1.29 | 0.25 | 0.26 | 0.05 | 0.99 | [-0.26, 0.77] |
| EDH Yes | 0.39 | -0.94 | 0.26 | 0.35 | 0.93 | [-1.48, -0.40] | 0.42 | -0.86 | 0.26 | 0.37 | 0.93 | [-1.38, -0.33] |
| Glucose | 1.04 | 0.04 | 0.05 | 0.07 | 0.99 | [-0.06, 0.15] | 1.06 | 0.06 | 0.02 | 0.22 | 0.96 | [0.01, 0.10] |
| Hemoglobin | 1.01 | 0.01 | 0.04 | 0.25 | 0.95 | [-0.07, 0.10] | 1 | 0 | 0.04 | 0.3 | 0.94 | [-0.08, 0.08] |
| Hypotension Yes | 1.41 | 0.34 | 0.27 | 0.01 | 1 | [-0.20, 0.88] | 1.43 | 0.36 | 0.22 | 0.11 | 0.98 | [-0.07, 0.79] |
| Hypoxia Yes | 1.4 | 0.34 | 0.28 | 0.07 | 0.99 | [-0.21, 0.88] | 1.44 | 0.36 | 0.2 | 0.09 | 0.98 | [-0.04, 0.77] |
| Marshall I | 1 | 0 | 0.62 | 0.41 | 0.92 | [-1.27, 1.27] | 1.16 | 0.14 | 0.6 | 0.43 | 0.92 | [-1.09, 1.38] |
| Marshall III/IV | 5.19 | 1.65 | 0.27 | 0.18 | 0.96 | [1.11, 2.19] | 5.83 | 1.76 | 0.25 | 0.12 | 0.98 | [1.26, 2.26] |
| Marshall V/VI | 3.97 | 1.38 | 0.24 | 0.5 | 0.91 | [0.87, 1.89] | 3.86 | 1.35 | 0.24 | 0.51 | 0.91 | [0.85, 1.85] |
| Pupils One | 2.67 | 0.98 | 0.44 | 0.15 | 0.97 | [0.11, 1.86] | 2.4 | 0.87 | 0.24 | 0.06 | 0.99 | [0.39, 1.35] |
| Pupils None | 5.98 | 1.79 | 0.2 | 0.06 | 0.99 | [1.40, 2.18] | 5.26 | 1.66 | 0.19 | 0.09 | 0.98 | [1.28, 2.04] |
| TSAH Yes | 1.49 | 0.4 | 0.3 | 0.34 | 0.94 | [-0.21, 1.00] | 1.59 | 0.47 | 0.28 | 0.27 | 0.95 | [-0.09, 1.03] |

**eTable 14.** LAB Unfavorable outcome Study-level model parameters. OR = Odds Ratio, *β =* Model coefficient, SE = Standard Error, FMI = Fraction of missing information, RE = relative efficiency due to missingness, 95% CI = 95% confidence interval of the coefficient

| **TRACK-TBI LAB Unfavorable** | | | | | | | **CENTER-TBI LAB Unfavorable** | | | | | |
| --- | --- | --- | --- | --- | --- | --- | --- | --- | --- | --- | --- | --- |
| Term | OR | *β* | *SE* | FMI | RE | 95% CI | OR | *β* | *SE* | FMI | RE | 95% CI |
| Age | 1.05 | 0.05 | 0.01 | 0.09 | 0.98 | [0.03, 0.07] | 1.03 | 0.03 | 0 | 0.08 | 0.98 | [0.02, 0.04] |
| GCS motor 1 | 2.27 | 0.82 | 0.29 | 0.03 | 0.99 | [0.25, 1.40] | 2.22 | 0.8 | 0.17 | 0.01 | 1 | [0.46, 1.13] |
| GCS motor 2 | 4.23 | 1.44 | 0.55 | 0.07 | 0.99 | [0.35, 2.53] | 4.24 | 1.45 | 0.38 | 0.03 | 0.99 | [0.71, 2.18] |
| GCS motor 3 | 1.84 | 0.61 | 0.68 | 0.06 | 0.99 | [-0.73, 1.95] | 2.3 | 0.83 | 0.32 | 0 | 1 | [0.20, 1.46] |
| GCS motor 4 | 1.14 | 0.13 | 0.38 | 0.06 | 0.99 | [-0.63, 0.88] | 1.53 | 0.43 | 0.25 | 0.01 | 1 | [-0.06, 0.91] |
| EDH Yes | 0.53 | -0.63 | 0.34 | 0.15 | 0.97 | [-1.30, 0.04] | 0.44 | -0.83 | 0.21 | 0.16 | 0.97 | [-1.23, -0.42] |
| Glucose | 1 | 0 | 0.04 | 0.2 | 0.96 | [-0.08, 0.08] | 1.12 | 0.11 | 0.04 | 0.69 | 0.88 | [0.01, 0.21] |
| Hemoglobin | 0.87 | -0.14 | 0.08 | 0.37 | 0.93 | [-0.30, 0.02] | 0.97 | -0.03 | 0.06 | 0.7 | 0.88 | [-0.16, 0.10] |
| Hypotension Yes | 2.28 | 0.82 | 0.39 | 0.01 | 1 | [0.05, 1.59] | 1.73 | 0.55 | 0.23 | 0.13 | 0.98 | [0.10, 1.00] |
| Hypoxia Yes | 1.64 | 0.5 | 0.35 | 0.03 | 0.99 | [-0.19, 1.18] | 0.91 | -0.1 | 0.22 | 0.13 | 0.98 | [-0.52, 0.33] |
| Marshall I | 0.33 | -1.12 | 1.11 | 0.39 | 0.93 | [-3.39, 1.16] | 0.62 | -0.47 | 0.43 | 0.39 | 0.93 | [-1.36, 0.42] |
| Marshall III/IV | 1.81 | 0.59 | 0.41 | 0.03 | 0.99 | [-0.20, 1.39] | 2.39 | 0.87 | 0.27 | 0.2 | 0.96 | [0.34, 1.40] |
| Marshall V/VI | 2.68 | 0.98 | 0.28 | 0.09 | 0.98 | [0.42, 1.54] | 2.4 | 0.87 | 0.2 | 0.34 | 0.94 | [0.48, 1.27] |
| Pupils One | 2.18 | 0.78 | 0.46 | 0.07 | 0.99 | [-0.12, 1.68] | 1.7 | 0.53 | 0.26 | 0.05 | 0.99 | [0.02, 1.04] |
| Pupils None | 2.92 | 1.07 | 0.36 | 0.22 | 0.96 | [0.36, 1.79] | 2.86 | 1.05 | 0.22 | 0.06 | 0.99 | [0.61, 1.49] |
| TSAH Yes | 2.53 | 0.93 | 0.35 | 0.01 | 1 | [0.23, 1.62] | 1.32 | 0.28 | 0.25 | 0.31 | 0.94 | [-0.22, 0.77] |

**eTable 15.** LAB Unfavorable outcome federated data model parameters. OR = Odds Ratio, *β =* Model coefficient, SE = Standard Error, FMI = Fraction of missing information, RE = relative efficiency due to missingness, 95% CI = 95% confidence interval of the coefficient

| **SLMA LAB Unfavorable** | | | | | | | **IRLS LAB Unfavorable** | | | | | |
| --- | --- | --- | --- | --- | --- | --- | --- | --- | --- | --- | --- | --- |
| Term | OR | *β* | *SE* | FMI | RE | 95% CI | OR | *β* | *SE* | FMI | RE | 95% CI |
| Age | 1.04 | 0.04 | 0 | 0.12 | 0.98 | [0.03, 0.05] | 1.04 | 0.04 | 0 | 0.06 | 0.99 | [0.03, 0.04] |
| GCS motor 1 | 2.23 | 0.8 | 0.15 | 0.02 | 1 | [0.51, 1.09] | 2.22 | 0.8 | 0.15 | 0.02 | 1 | [0.51, 1.08] |
| GCS motor 2 | 4.24 | 1.44 | 0.31 | 0.04 | 0.99 | [0.84, 2.05] | 4.11 | 1.41 | 0.31 | 0.04 | 0.99 | [0.81, 2.01] |
| GCS motor 3 | 2.2 | 0.79 | 0.29 | 0.02 | 1 | [0.22, 1.36] | 2.13 | 0.75 | 0.28 | 0.02 | 1 | [0.20, 1.31] |
| GCS motor 4 | 1.4 | 0.34 | 0.21 | 0.04 | 0.99 | [-0.07, 0.74] | 1.39 | 0.33 | 0.21 | 0.04 | 0.99 | [-0.07, 0.73] |
| EDH Yes | 0.46 | -0.77 | 0.18 | 0.15 | 0.97 | [-1.12, -0.43] | 0.46 | -0.78 | 0.17 | 0.16 | 0.97 | [-1.12, -0.43] |
| Glucose | 1.06 | 0.06 | 0.05 | 0.19 | 0.96 | [-0.03, 0.15] | 1.08 | 0.07 | 0.03 | 0.56 | 0.9 | [0.01, 0.14] |
| Hemoglobin | 0.94 | -0.06 | 0.05 | 0.47 | 0.91 | [-0.17, 0.04] | 0.95 | -0.05 | 0.05 | 0.72 | 0.87 | [-0.17, 0.06] |
| Hypotension Yes | 1.85 | 0.61 | 0.2 | 0.09 | 0.98 | [0.23, 1.00] | 1.85 | 0.61 | 0.19 | 0.07 | 0.99 | [0.24, 0.98] |
| Hypoxia Yes | 1.07 | 0.06 | 0.19 | 0.09 | 0.98 | [-0.31, 0.44] | 1.05 | 0.05 | 0.18 | 0.07 | 0.99 | [-0.30, 0.40] |
| Marshall I | 0.58 | -0.54 | 0.39 | 0.34 | 0.94 | [-1.32, 0.24] | 0.57 | -0.57 | 0.36 | 0.29 | 0.95 | [-1.29, 0.16] |
| Marshall III/IV | 2.22 | 0.8 | 0.22 | 0.11 | 0.98 | [0.36, 1.23] | 2.24 | 0.81 | 0.21 | 0.1 | 0.98 | [0.39, 1.23] |
| Marshall V/VI | 2.47 | 0.9 | 0.17 | 0.33 | 0.94 | [0.56, 1.24] | 2.45 | 0.89 | 0.16 | 0.33 | 0.94 | [0.56, 1.23] |
| Pupils One | 1.8 | 0.59 | 0.23 | 0.06 | 0.99 | [0.14, 1.04] | 1.88 | 0.63 | 0.23 | 0.06 | 0.99 | [0.19, 1.08] |
| Pupils None | 2.88 | 1.06 | 0.19 | 0.14 | 0.97 | [0.68, 1.44] | 2.77 | 1.02 | 0.19 | 0.13 | 0.98 | [0.65, 1.39] |
| TSAH Yes | 1.6 | 0.47 | 0.22 | 0.15 | 0.97 | [0.03, 0.91] | 1.57 | 0.45 | 0.19 | 0.18 | 0.96 | [0.07, 0.83] |
